# Emergence of a multidrug resistant and virulent *Streptococcus pneumoniae* lineage mediates serotype replacement after PCV13

**DOI:** 10.1101/2021.11.24.21266813

**Authors:** Stephanie W Lo, Kate Mellor, Robert Cohen, Alba Redin Alonso, Sophie Belman, Narender Kumar, Paulina A Hawkins, Rebecca A Gladstone, Anne von Gottberg, Balaji Veeraraghavan, KL Ravikumar, Rama Kandasamy, Andrew J Pollard, Samir K Saha, Godfrey Bigogo, Martin Antonio, Brenda Kwambana-Adams, Shaper Mirza, Sadia Shakoor, Imran Nisar, Jennifer E Cornick, Deborah Lehmann, Rebecca L Ford, Betuel Sigauque, Paul Turner, Jennifer Moïsi, Stephen K Obaro, Ron Dagan, Naima Elmdaghri, Anna Skoczyńska, Hui Wang, Philip E Carter, Keith P Klugman, Gail Rodgers, Robert F Breiman, Lesley McGee, Stephen D Bentley, Carmen Muñoz Almagro, Emmanuelle Varon, The Global Pneumococcal Sequencing Consortium

**Affiliations:** Parasites and Microbes, Wellcome Sanger Institute, Hinxton, UK; ACTIV, Association Clinique et Thérapeutique Infantile du Val-de-Marne, Saint Maur- des-Fossés, France; GPIP, Groupe de Pathologie Infectieuse Pédiatrique, Paris, France; AFPA, Association Française de Pédiatrie Ambulatoire, Saint-Germain-en-Laye, France; Université Paris Est, IMRB-GRC GEMINI, Créteil, France; Clinical Research Center (CRC), Centre Hospitalier Intercommunal de Créteil, Créteil, Créteil, France; Unité Court Séjour, Petits nourrissons, Service de Néonatalogie, Centre Hospitalier Intercommunal de Créteil, France; Department of Molecular Microbiology, Hospital Sant Joan de Deu, Barcelona, Spain; School of Medicine, Universitat Internacional de Catalunya, Barcelona, Spain; Spanish Network of Epidemiology and Public Health, CIBERESP, Instituto de Salud Carlos III, Madrid, Spain; Rollins School Public Health, Emory University, Atlanta, GA, USA; Department of Biostatistics, Institute of Clinical Medicine, Faculty of Medicine, University of Oslo, Oslo, Norway; Centre for Respiratory Diseases and Meningitis, National Institute for Communicable Diseases, Johannesburg, South Africa; Department of Clinical Microbiology, Christian Medical College, Vellore, Tamil Nadu 632004 India; Central Research Laboratory, KIMS Hospital and Research Centre, KR Road, VV Purum, Bangalore, Karnataka 560 004 India; Oxford Vaccine Group, Department of Paediatrics, University of Oxford, Churchill Hospital, Oxford OX3 7LE, UK NIHR Oxford Biomedical Research Centre, Oxford, OX3 7LE, UK; University of New South Wales, School of Women and Children’s Health, University of New South Wales, Sydney, Australia; Child Health Research Foundation, Dhaka, Bangladesh; Kenya Medical Research Institute, Kisumu, Kenya; WHO Collaborating Centre for New Vaccines Surveillance, Medical Research Council Unit The Gambia at The London School of Hygiene & Tropical Medicine, Fajara, The Gambia; NIHR Global Health Research Unit on Mucosal Pathogens, Division of Infection and Immunity, University College London, London, UK; Microbiology and Immunology Laboratory, Department of Biology, Lahore University of Management Sciences, Lahore, Pakistan; Department of Pediatrics and Child Health, Aga Khan University, Stadium Road, Karachi, 74800 Pakistan; Malawi-Liverpool-Wellcome-Trust, Blantyre, Malawi; Wesfarmers Centre of Vaccines and Infectious Diseases, Telethon Kids Institute, The University of Western Australia, Perth, Australia; Papua New Guinea Institute of Medical Research, Goroka, Papua New Guinea; Centro de Investigação em Saúde da Manhiça (CISM), Maputo, Mozambique; Cambodia Oxford Medical Research Unit, Angkor Hospital for Children, Siem Reap; Centre for Tropical Medicine and Global Health, Nuffield Department of Medicine, University of Oxford, United Kingdom; Agence de Médecine Préventive, Abidjan, Cote d’Ivoire; Division of Pediatric Infectious Disease, University of Nebraska Medical Center Omaha, Omaha, NE, USA; International Foundation against Infectious Diseases in Nigeria, Abuja, Nigeria; Ben-Gurion University of the Negev, Beer-Sheva, Israel; Department of Microbiology, Faculty of Medicine and Pharmacy of Casablanca, Hassan II University of Casablanca, Casablanca, Morocco; Bacteriology-Virology and Hospital Hygiene Laboratory, Ibn Rochd University Hospital Centre, Casablanca, Morocco; Department of Epidemiology and Clinical Microbiology, National Medicines Institute, Chełmska 30/34, 00-725, Warsaw, Poland; Peking University People ’s Hospital, China; Institute of Environmental Science and Research Limited, Kenepuru Science Centre, Porirua, New Zealand; Pneumonia Program, Bill & Melinda Gates Foundation, Seattle, WA 98119, USA; Centers for Disease Control and Prevention, Atlanta, GA, USA; Emory Global Health Institute, Emory University, Atlanta, GA, USA; National Reference Center for Pneumococci, Centre Hospitalier Intercommunal de Créteil, Créteil, France

## Abstract

**Background:** Pneumococcal Conjugate Vaccine (PCV) which targets up to 13 serotypes of *Streptococcus pneumoniae* is very effective at reducing disease in young children; however, rapid increases in replacement with non-PCV serotypes remains a concern. Serotype 24F is one of the major invasive serotypes that mediates serotype replacement in France and multiple other countries. We aimed to identify the major pneumococcal lineage that has driven the increase of serotype 24F in France, and provide context for the findings by investigating the global diversity of serotype 24F pneumococci and characterise the driver lineage from a global perspective and elucidate its spatiotemporal transmission in France and across the world.

**Methods:** We whole-genome sequenced a collection of 419 serotype 24F *S. pneumoniae* from asymptomatic carriers and invasive disease cases among individuals <18 years old in France between 2003 and 2018. Genomes were clustered into Global Pneumococcal Sequence Clusters (GPSCs) and clonal complexes (CCs) so as to identify the lineages that drove the increase in serotype 24F in France. For each serotype 24F lineage, we evaluated the invasive disease potential and propensity to cause meningitis by comparing the proportion of invasive disease cases with that of carriers. To provide a global context of serotype 24F and the driver lineage, we extracted relevant genomes and metadata from the Global Pneumococcal Sequencing (GPS) project database (n=25,590) and additionally sequenced a collection of 91 pneumococcal isolates belonging to the lineage that were responsible for the serotype 24F increase in Spain during the PCV introduction for comparison. Phylogenetic, evolutionary, and spatiotemporal analysis were conducted to understand the mechanism underlying the global spread of serotype 24F, evolutionary history and long-range transmissions of the driver lineage.

**Findings:** A multidrug-resistant pneumococcal lineage GPSC10 (CC230) drove the serotype 24F increase in both carriage and invasive disease in France after PCV13 introduction. When compared with other serotype 24F lineages, it exhibited a 1.4-fold higher invasive disease potential and 1.6-fold higher propensity to cause meningitis. Globally, serotype 24F was widespread, largely due to clonal dissemination of GPSC10, GPSC16 (CC66) and GPSC206 (CC7701) rather than recent capsular switching. Among these lineages, only GPSC10 was multidrug-resistant. It expressed 17 serotypes, with only 6 included in PCV13 and none of the expected PCVs covered all serotypes expressed by this lineage. Global phylogeny of GPSC10 showed that all serotype 24F isolates except for one were clustered together, regardless of its country of origin. Long-range transmissions of GPSC10-24F from Europe to Israel, Morocco and India were detected. Spatiotemporal analysis revealed that it took ∼5 years for GPSC10- 24F to spread across French provinces. In Spain, we detected that the serotype 24F driver lineage GPSC10 underwent a rapid change of serotype composition from serotype 19A to 24F during the introduction of PCV13 (targets 19A but not 24F), indicating that pre-existence of serotype variants enabled GPSC10 to survive and expand under vaccine-selective pressure.

**Interpretation:** Our work further shows the utility of bacterial genome sequencing to better understand the pneumococcal lineages behind the serotype changes and reveals that GPSC10 alone is a challenge for serotype-based vaccine strategy. More systematic investigation to identify lineages like GPSC10 will better inform and improve next-generation preventive strategies against pneumococcal diseases.

**Funding:** Bill & Melinda Gates Foundation, Wellcome Sanger Institute, and the US Centers for Disease Control.

## Introduction

Pneumococcal conjugate vaccines (PCV), which targets up to 13 capsule serotypes of *Streptococcus pneumoniae* that account for most of the diseases in infants, have been very effective at reducing disease worldwide.^1^ However, increases in replacement with non-PCV serotypes remain a concern,^2–6^ such as serotype 19A after PCV7 in the USA.^7^ In France, a sharp increase in pneumococcal meningitis cases occurred five years after the roll out of PCV13, mainly driven by a non-PCV13 serotype 24F.^8, 9^ This serotype also mediated serotype replacement in multiple other countries such as Argentina^10^, Canada,^11^ Denmark,^12^ Germany,^13^ Israel,^14^ Italy,^15^ Japan,^16^ Lebanon,^17^ Norway,^18^ Spain,^19, 20^ and UK^21^, and was reported to be the predominant serotype causing invasive pneumococcal disease (IPD) in Portugal^22^ after PCV13 introduction.

Compared with most of the non-PCV13 serotypes, the serotype 24F capsule has a high invasive disease potential^23^ and propensity to cause meningitis.^24^ In France, the fatality rate for meningitis due to serotype 24F pneumococci was 13%, similar to that (11%) caused by pneumococci expressing other serotypes.^8^ In some countries, the serotype 24F increase was concomitant with increasing prevalence of penicillin resistance in IPD overall^8, 15^ and IPDs due to non-vaccine serotypes.^10^ Despite serotype 24F being an important emerging serotype with high invasiveness and potential for antimicrobial resistance, it is not included, to our knowledge, in any expected future PCV formulations (PCV15, 20, 24).

By delineating pneumococcal lineages into Global Pneumococcal Sequence Clusters (GPSCs, aka lineages) using variations across the entire genome^25^ and/or clonal complexes (CCs) defined by nucleotide sequences of seven housekeeping genes,^26^ we observed that pneumococcal lineages driving the increase in serotype 24F varied between countries. For example, the increase was mainly driven by GPSC10 (CC230) in Argentina,^10^ Lebanon^17^ and Spain^27^, by GPSC6 (CC156) in Denmark^12^, by GPSC106 (CC2572) in Japan.^16^ Here we applied whole genome sequencing to investigate the pneumococcal lineages driving the increase in serotype 24F after PCV13 introduction in France. Merging with >25,000 pneumococcal genomes from 56 countries in the Global Pneumococcal Sequencing (GPS) project database, we provide context for the findings from France by investigating the global diversity of serotype 24F pneumococci. We characterise the driver lineage from a global perspective and elucidate its spatiotemporal transmission in France and across the world.

## Methods

### Study design

A representative set of French serotype 24F pneumococcal isolates collected through a nationwide hospital-based active surveillance for IPD^8, 9^ and carriage survey^28^ across the country were whole-genome sequenced. The collection included isolates from invasive disease cases (n=190) and asymptomatic colonisation (n=229) among individuals <18 years old between 2003 and 2018 (Figure S1). The study period spanned across different phases of PCV introduction: from PCV7 use in target groups of children (e.g. children in a daycare) to generalised use of PCV13 for all children and the subsequent increase of 24F in 2015. To identify the pneumococcal lineage(s) driving the increase in 24F, we grouped isolates into GPSCs and CCs, and evaluated each lineage’s invasive disease potential and propensity to cause meningitis by calculating odds ratio by reference to carriage.^29^

We then contextualised the serotype 24F and the driver lineage (GPSC10) from a global perspective by including additional genomes from the GPS project database (n=25,590 from 55 countries, last accessed on 2^nd^ October 2021). We additionally sequenced a collection of 91 pneumococcal isolates that were responsible for the serotype 24F increase in Spain for comparison. These isolates were collected from children aged <5 years old in the Catalan support laboratory for non-mandatory molecular surveillance of IPD located at Hospital Sant Joan de Déu, Barcelona between 2009 and 2018.^27^ To understand the genetic diversity, phylogenetic analysis was carried out on 642 serotype 24F isolates from 29 countries across six continents (Africa, Asia, Australia, Europe, North America, and Latin America). An international collection of 888 GPSC10 isolates from 33 countries, regardless of serotype, were included for phylogenetic, evolutionary, and spatiotemporal analysis.

### Sequencing and genomic characterisation

The pneumococcal isolates in this study were whole-genome sequenced at the Wellcome Sanger Institute (Hinxton, UK) on an Illumina HiSeq sequencer. The sequence reads were subjected to quality control based on the criteria as previously described^25^. We characterised each genome by assigning GPSC using PopPUNK^25, 30^, sequence type (ST) by MLSTcheck^31^ and then grouped STs into CC using Eburst^32^, predicted serotypes using SeroBA^33^, and resistance profile of 17 antibiotics, including penicillin, chloramphenicol, erythromycin, cotrimoxazole and tetracycline, using a pipeline developed by the Streptococcus Laboratory at the Centers for Disease Control and Prevention Atlanta, USA.^33, 34^ Multidrug resistance (MDR) was defined as an isolate resistant to ≥3 antibiotic classes. In our previous large-scale analysis, we showed a high concordance between GPSC and CC, therefore sequence type (ST) identified in previous studies were used to infer GPSC in this study. All sequencing reads were deposited in European Nucleotide Archive (ENA) and the accession number is enclosed with the metadata and in silico output in the supplementary file 1. Additional details on DNA extraction, genome quality control criteria, de novo assembly, annotation and in silico serotyping within serogroup 24 were described in supplementary file 2.

### Phylogenetic analysis

We performed phylogenetic analysis on all serotype 24F isolates by constructing a maximum likelihood tree using FastTree version 2.1.10 with the general time reversible substitution model.^34^ Phylogenies were built based on single nucleotide polymorphisms (SNPs) extracted from individual alignment generated by mapping reads to a reference genome of *S. pneumoniae* ATCC 700669 (NCBI accession number FM211187) using SMALT version 0.7.4, with default settings^35^ and to a reference sequence of serotype 24F capsular encoding region (i.e. *cps*, NCBI accession number CR931688) using Burrows Wheeler Aligner (BWA) version 0.7.17-r1188^36^. The phylogenetic trees were then overlaid with epidemiological data and *in silico* output as described above and visualised in Microreact^37^ at https://microreact.org/project/global_24F and https://microreact.org/project/global_24F_cps, respectively.

### Evolutionary and spatiotemporal analysis

Sequence reads of GPSC10 (CC230) isolates were mapped to the GPSC10 reference genome Denmark^14^-32 (ENA accession number ERS1706837) using BWA version 0.7.17- r1188.^36^ Recombination was detected and removed using GUBBINS version 2.4.1^38^. A recombination-free phylogeny was produced with RAxML version 8.2.8^39^ and then visualised in Microreact^37^, together with metadata (https://microreact.org/project/global_GPSC10). Bayesian phylogenetic analysis on a subset of serotype 24F isolates within GPSC10 was conducted to generate a time-resolved phylogeny and provide estimates of the median effective population size over time with a 95% highest posterior density to detect any exponential increase in population using BEAST Bayesian skyline model version 2.6.3^40^. The time-resolved phylogeny can be visualised at https://microreact.org/project/GPSC10-24F.

To calculate time taken for GPSC10-24F to spread across France, we inferred the evolutionary time between all pairs of genomes from the time-resolved phylogeny. We then utilized an risk ratio framework to calculate the odds that a pair of genomes from within the same province (0-50km) compared with pairs isolated from locations between 250-350km apart (the mean distance between different French provinces is 330km) had a specified time-to-most-recent-common-ancestor (tMRCA) across rolling 2 year divergence time windows from 2 to 20 years.^41^ We restricted the analysis to pairs isolated within the same year to mitigate variable geographic sampling across years. A risk ratio close to one indicated an equal chance that a pair of isolates diverged from the same tMRCA within and between provinces. To determine uncertainty, we ran this with 100 bootstrapped iterations sampling with replacement. We determined the significance of the relationship between the tMRCA and risk ratio using a generalized linear mixed model. The spatiotemporal analysis was run using R version 3.6.0.

### Statistical analysis

To determine if differences in proportions between two groups were significant, two-sided Fisher’s exact test was used. Two-sided p values of less than 0.05 were considered significant. Multiple testing correction was carried out using the Benjamini-Hochberg false discovery rate of 5%.^42^ We grouped the French isolates into four vaccine periods as previously described by Ouldali et al.^8^: 1) targeted PCV7 period (2003-2005) in which PCV7 only reimbursed and recommended for children in a day-care centre with ≥2 other children, children in families with >2 children, or children breastfed for fewer than 2 months; 2) generalised PCV7 period (2006- 2010) in which PCV7 were applied to all children younger than 2 years; 3) early PCV13 period (2011-2014) and 4) late PCV13 period (2015-2018) in which PCV7 was replaced with PCV13, without catch up. Using these four periods, we compared the prevalence of GPSCs. Statistical tests were performed in R version 3.6.0.

## Results

The 419 French serotype 24F pneumococcal isolates were sequenced and all passed sequencing quality control criteria. The collection was delineated mainly to 3 lineages GPSC10 (CC230, 41.5%), GPSC6 (CC156, 38.2%), and GPSC16 (CC66, 17.7%) and a small percentage of isolates were other lineages: GPSC44 (CC177, 1.4%), GPSC18 (CC15, 0.7%) and GPSC5 (CC172, 0.5%). Over the study period, clonal replacement was observed in overall disease, meningitis and carriage isolates (Figure 1). Overall, GPSC16 showed a significant decrease since the generalised PCV7 period (2006-2010). In contrast, GPSC6 significantly increased from 9.9% in the generalised PCV7 period to 62.7% in the early PCV13 period (2011-2014). Whilst GPSC6 decreased to 30.2% in the late PCV13 period (2015-2018), GPSC10 significantly increased from 23% in the early PCV13 period to 65.1% in the late PCV13 period (p value <0.0001 for all changes, Table S1). In the late PCV13 period, GPSC10 became the predominant lineage, accounting for 100% (10/10) of the pneumonia cases caused by serotype 24F pneumococcus, 74% (35/47) of meningitis cases, 67% (4/6) of other infections, 50% (11/22) of bacteraemias and 60% (50/84) of asymptomatic colonisation. In contrast to the pan-susceptible GPSC16, the replacement lineages GPSC6 and GPSC10 were cotrimoxazole-resistant and MDR, respectively.

**Figure 1.**
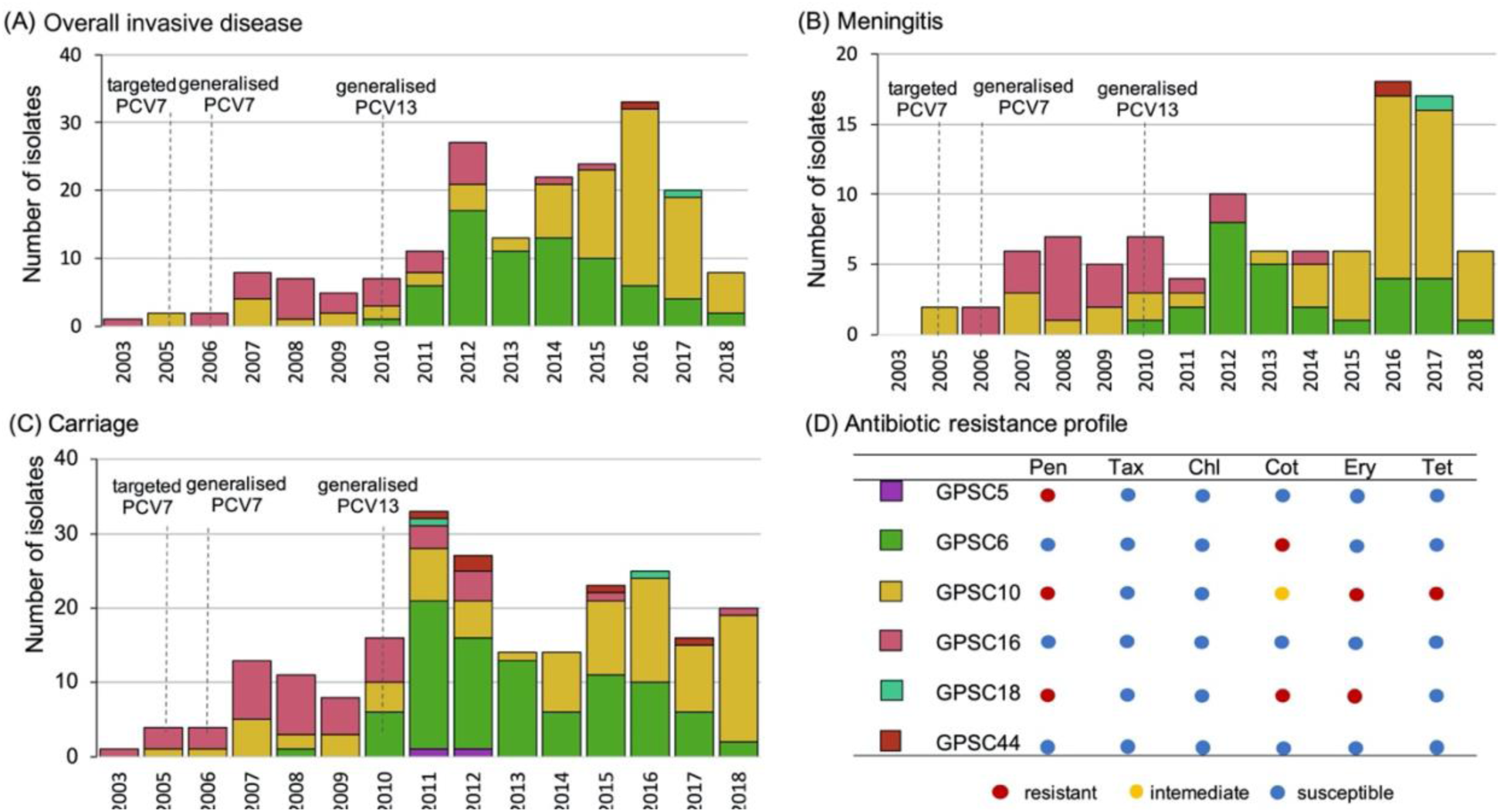
(A-C) Proportion of serotype 24F pneumococcal lineages from France between 2003 and 2018 and (D) predominant antibiotic resistance profiles of each lineage. GPSC, global pneumococcal sequence cluster; PCV, pneumococcal conjugate vaccine; Pen, penicillin; Tax, cefotaxime; Chl, chloramphenicol; Ery, erythromycin; Tet, tetracycline.

Compared with other serotype 24F lineages, only GPSC10 was more frequently detected in overall invasive disease (OR:1.38, 95%CI: 0.93-2.04, p=0.107) and meningitis (OR:1.57, 95% CI:0.98-2.51, p=0.06) cases than in asymptomatic colonisation. Although the finding did not reach statistical significance, it suggested that this lineage had relatively high invasive disease potential and propensity to cause meningitis (Figure S2 and Table S2). In contrast, GPSC6 was more frequently identified in carriage than in meningitis isolates (OR:0.58, 95%CI:0.35- 0.97, p=0.038).

The global collection of 642 serotype 24F isolates revealed that this serotype was widely distributed across 29 countries (Figure S3). We delineated the global collection of 642 serotype 24F isolates into 20 GPSCs. GPSC10, 16, 150 and 206 were the most common lineages, accounting for 68% (439/642) of the overall GPS collection and 78% (96/123) of a sub-collection includes isolates randomly selected from disease surveillance systems and carriage surveys from 21 countries (Figure S3). GPSC10, 16 and 206 are globally-spreading lineages, while GPSC150 is only detected in West Africa, except for three isolates from Israel. Among these most common lineages, only GPSC10 was MDR with a majority of isolates exhibiting resistance to penicillin, cotrimoxazole, erythromycin and tetracycline (Figure S4).

Phylogenetic analysis of the serotype 24F *cps* revealed a strong clonal but not geographical structure (Figure S5). The *cps* belonging to the same GPSC were clustered together regardless of the isolates’ country of origin. This finding indicated that after a single capsular switching to 24F, the serotype variants clonally disseminated across different geographical regions majorly at the genetic background of GPSC10, 16 and 206. Only a few capsular switching events were observed based on the high similarity in *cps* between lineages. For example, three GPSC18 isolates from France share highly similar *cps* with GPSC10, suggesting a capsular switching between GPSC10 and GPSC18. GPSC18 expressing serotype 24F was not detected elsewhere but in France according to two largest pneumococcal isolate databases, the GPS project and pubMLST database (last accessed on 12^th^ August 2021). GPSC18-24F was first detected in 2011 in this collection of isolates, a year after the introduction of PCV13 in France, without significant expansion (Figure 1). GPSC18 was a MDR lineage exhibiting resistance to penicillin, cotrimoxazole and erythromycin and found to cause both invasive disease and asymptomatic colonisation.

Of all serotype 24F lineages, GPSC10 was the only one detected with high invasive disease potential and multidrug resistance. It was responsible for the increase of serotype 24F in France, and one of the major lineages mediating the global spread of serotype 24F. We further investigated this lineage from a global perspective using an international collection of 888 GPSC10 isolates. This lineage was detected in 33 different countries across Africa, Asia, Europe, North and South America (Figure S6). It expressed 17 different serotypes, including 3, 6A, 6C, 7B, 10A, 11A, 13, 14, 15B, 15C, 17F, 19A, 19F, 23A, 23B, 23F and 24F. Only 6 of them (serotypes 3, 6A, 14, 19A, 19F and 23F) were covered by the current PCVs (PCV10/13) that were approved for children use. An additional 3 serotypes (10A, 11A 15B) are covered by Pfizer’s planned 20-valent vaccine, and 4 (10A, 11A 15B, 17F) by Merck’s planned 24- valent vaccine. The latter two were not approved for children use at the time of writing. At present, not a single vaccine is known to cover all 17 serotypes expressed by GPSC10. Concerningly, GPSC10 is among the top 5 lineages in India,^43^ Pakistan and Nepal (Table S4), where pneumococcal disease burden is highest.^1^ In these three countries, 15 serotype variants were detected in GPSC10 and only 6 were included in PCV13, underlining its potential to cause serotype replacement in the future. Internationally, GPSC10 was consistently found to express multiple serotypes and be multidrug-resistant.

The global GPSC10 phylogeny showed that all serotype 24F isolates but one were clustered together, regardless of their country of origin (Figure 2A). This finding is consistent with the 24F *cps* phylogeny, which indicated the global dissemination of GPSC10-24F was largely mediated by clonal spread rather than capsular switching. We analysed a collection of 91 GPSC10 isolates from Spain and detected a rapid change in serotype composition from serotype 19A to 24F after the implementation of PCV13 in the target group of children (Figure S7). The 19A and 24F serotype variants from Spain were separated in long branches on the global phylogeny, further indicating that the capsular switch predates the vaccine introduction and coexistence of both GPSC10 serotype variants in Spain.

**Figure 2.**
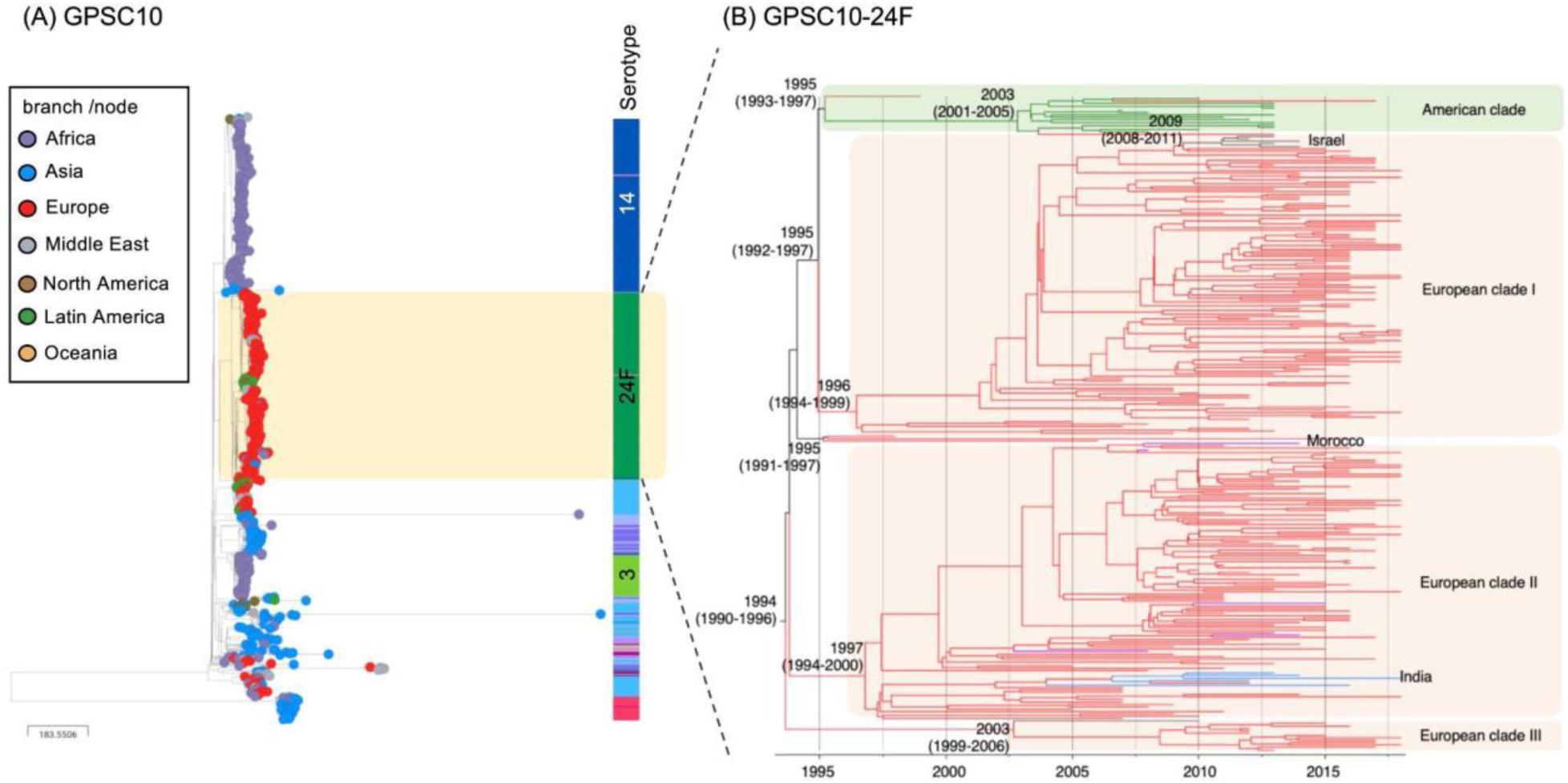
(A) Global phylogeny of GPSC10 and (B) time-resolved phylogeny of a cluster of GPSC10-24F *Streptococcus pneumoniae*. All but one serotype 24F isolates were clustered together, regardless of their country of origin, indicating a clonal spread of GPSC10-24F across the world. The GPSC10 global phylogeny can be interactively visualised at https://microreact.org/project/global_GPSC10/21948517 and time-resolved phylogeny at https://microreact.org/project/GPSC10-24F/8735223a

A time-resolved phylogeny was built on the cluster of 276 GPSC10-24F isolates and revealed four sub-clusters: EU-clade-I (dominated by ST4253), EU-clade-II (ST230 and ST4677), EU- clade-III (ST230) and American (ST230) clade (Figure 2B). EU-clade-I and -II were estimated to emerge around the 1990s and then clonally expanded. EU-clade-III was relatively small and estimated to emerge around late 1990s to early 2000s, with a majority of isolates from France and one from Qatar. Throughout the study period, EU-clade-I accounts for most (70%, 112/160) of the French GPSC10-24F isolates and drove the 24F increase while most of Spanish isolates (88%, 58/66) belonged to EU-clade II (Figure S8). In the EU-clade-I, long-range transmission was observed from Europe to Israel in the late 2000s. In the EU-clade-II, multiple transmissions from Europe to Morocco were detected and a single transmission from Europe to India in the early 2000s was followed by a clonal expansion.

We reconstructed the population dynamics over time on EU-clade-I and EU-clade-II using the Bayesian skyline model. The EU-clade-I was predicted to have three exponential increases in effective population size in around 1995, 2004 and 2013, while EU-clade-II had one in around 2010 (Figure 3A and 3B). The most recent increase in EU-clade-I coincided with the observed prevalence increase of GPSC10-24F in both overall disease and carriage isolates from France (Figure 3A, 3C-D). Spatiotemporal analysis of the 174 GPSC10-24F isolates from France indicated that a pair of pneumococcal isolates was more likely to be recovered within the same province if they were <5 years diverged from their MRCA (p=0.0083). Pairs which had diverged 5 or more years ago had a risk ratio stably surrounding 1 and no downward trend (p=0.4449). This suggests that the GPSC10-24F population was homogenous across French provinces after 4 years (Figure 4 and Table S3).

**Figure 3.**
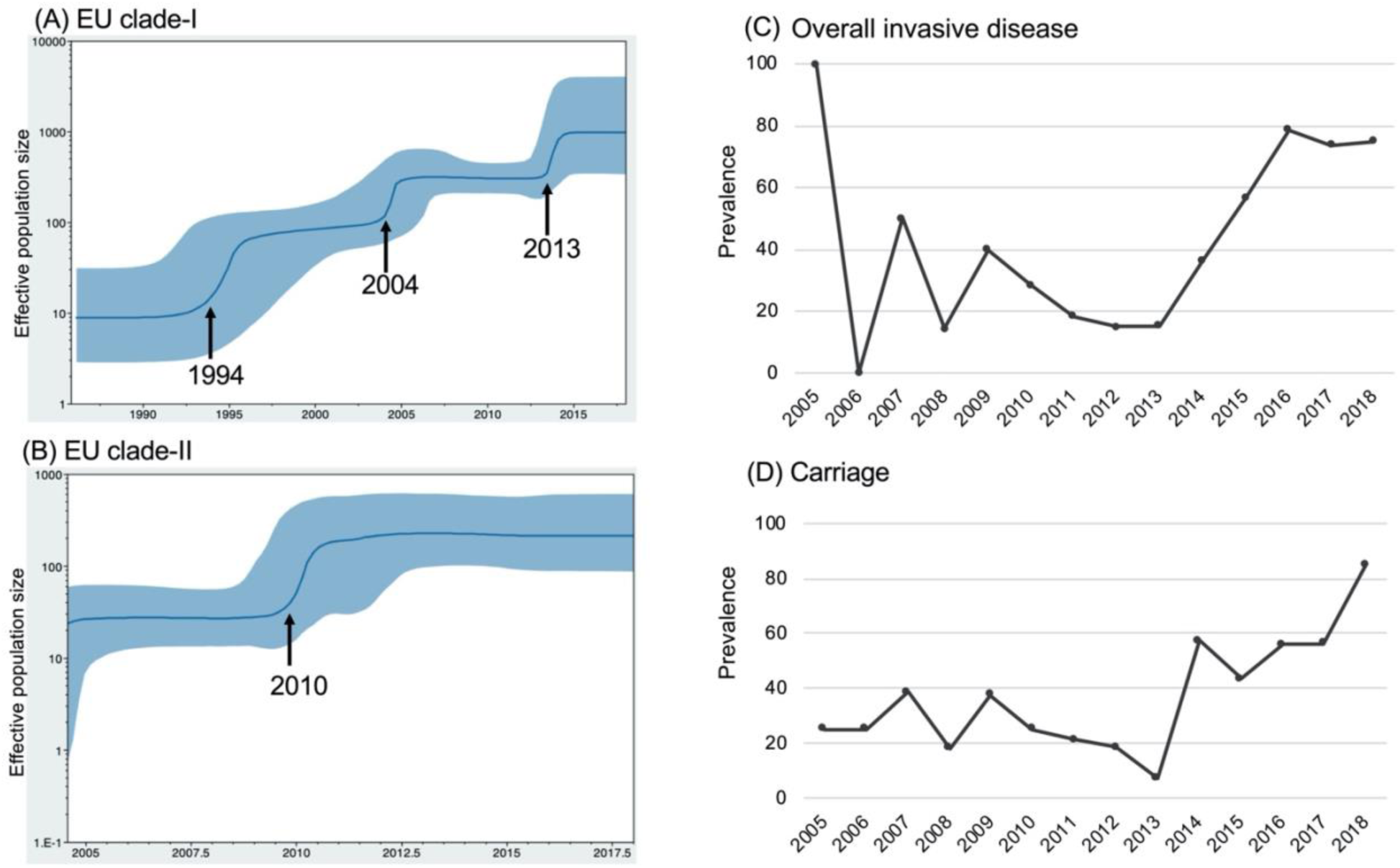
Bayesian skyline plots of estimated median effective population size of EU-clade-I and EU-clade II of GPSC10-24F *Streptococcus pneumoniae* over time (A and B), observed prevalence of GPSC10-24F *S. pneumoniae* from overall invasive disease and carriage in France over the collection year (C and D). The figure demonstrates three and one exponential increases in effective population size in EU-clade-I and EU-clade-II, respectively. The most recent increase in EU-clade-I coincided with the observed prevalence increase of GPSC10-24F in both overall disease and carriage isolates in France.

**Figure 4.**
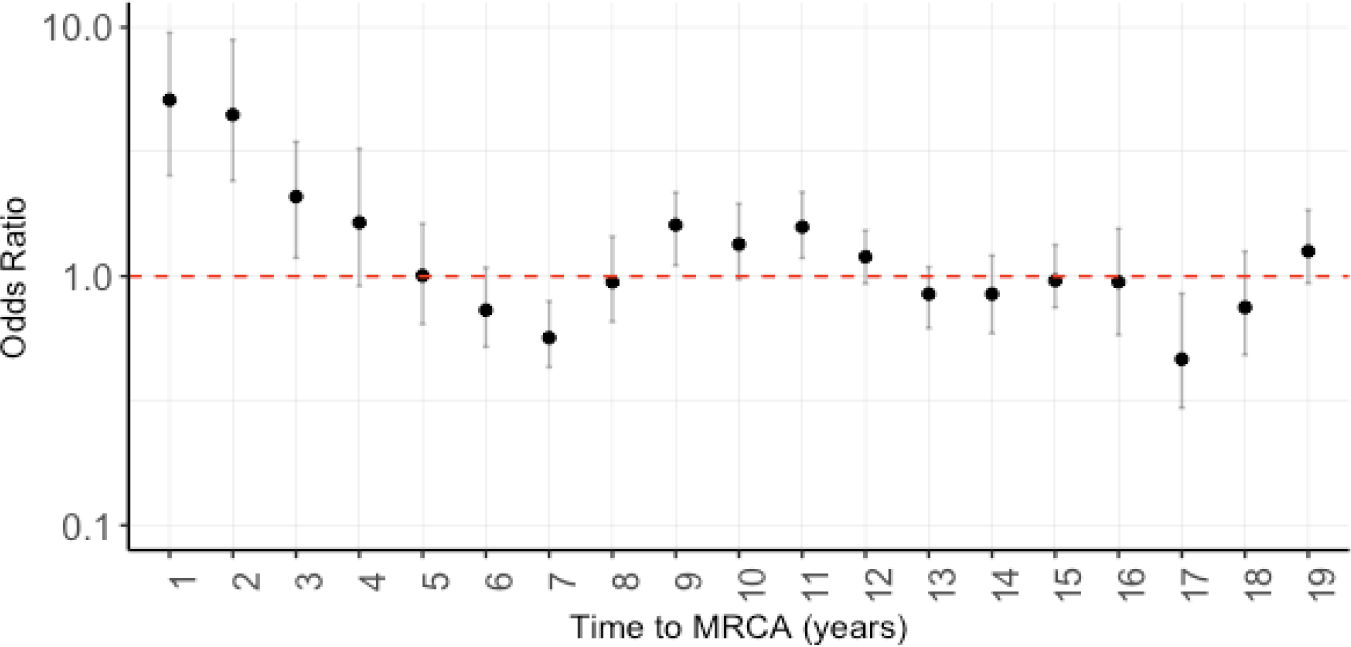
Spatiotemporal analysis of GPSC10-24F sub-lineage from France. A pairwise odds ratio was calculated for two samples being diverged from time-to-most-recent-common- ancestor (tMRCA) and recovered from the same French province. Odds ratio higher than one indicates that a pair of isolates is more likely to be recovered within the same French province. Pairs diverged 5 or more years ago had an odd ∼ 1 without up- or downward trend, indicating an equal chance to recover the sample pair within and between French provinces.

## Discussion

Our results showed the emergence of a virulent and MDR pneumococcal lineage GPSC10 that was responsible for the increase of invasive pneumococcal disease in France and the global spread of the invasive serotype 24F, which is not included in the current or expected PCVs. Due to its recombinogenic nature,^44^ this lineage is capable of simultaneously expressing a wide range of serotypes to facilitate its adaptation under the vaccine-selective pressure. Together with its transmissibility, GPSC10 should therefore be regarded as a high-risk lineage that could diminish the benefits of the vaccination programme worldwide over time.

Over the two decades since the advent of PCV, GPSC10 has mediated serotype replacement in multiple countries. After the introduction of PCV7/10, 19A became the predominant serotype causing invasive disease in Europe, such increase was partially driven by GPSC10, together with GPSC1 (CC320).^45–50^ In France and Spain, the major contributor of serotype 19A was GPSC10 in the post-PCV7 period.^50, 51^ The use of PCV13 (which targets 19A) effectively reduced serotype 19A but with a concurrent increase in serotype 24F in Europe.^8, 15, 22, 27, 52^ Using the Spanish collection, we observed a rapid change in serotype composition within GPSC10 from 19A to 24F after PCV13. A similar serotype change within GPSC10 was also observed in Argentina^10^ and Israel^53^ after PCV13, demonstrating that the pre-existence of serotype variants enables GPSC10 to survive and expand under the vaccine-selective pressure.

Serotype 24F pneumococci were infrequent causes of invasive disease in Europe in the 1980s^54^ and the earliest GPSC10 lineage was detected in Denmark 1996 expressing serotype 14.^55^ GPSC10-24F was first reported from three adult patients in Naples, Italy between 1997- 1998^56^; these findings coincide with our model prediction of the emergence of this clone in Europe. Since then, GPSC10-24F started to be more frequently detected in children^8, 45^ and adults^57, 58^ from Southern Europe. In spite of the geographical proximity between France and Spain, the increase in serotype 24F was mainly driven by two different clones, GPSC10 EU- clade-I and EU-clade-II, respectively. This finding may suggest trans-border transmission is present, but may not be as frequent as transmission within a country, resulting in evolution of two clones in parallel.

Although the global spread of serotype 24F is largely due to the clonal spread of three pneumococcal lineages (GPSC10, 16 and 206), lineage that drove the serotype 24F increase differs between countries. This variation could be partially explained by the differences in antibiotic-selective pressure. For example, the 24F driver lineage in Denmark was GPSC6.^12^ Unlike GPSC10, GPSC6 was susceptible to penicillin and erythromycin. This observation coincided with the lower consumption of penicillin and macrolide (a class of antibiotic includes erythromycin) in Denmark, as compared with other countries such as France^8, 9^, Lebanon^17^ and Spain^19, 20^ where serotype 24F increase was mediated by the multidrug-resistant GPSC10 (Table S5). The latter three countries consumed 1.8-2.2 times more penicillin and 1.2-2.0 times more macrolide than that in Denmark. GPSC10 also mediated the serotype 24F increase in Argentina^10^ where macrolide consumption was 1.2 times higher than Denmark, though penicillin consumption was almost similar. The high consumption of penicillin and/or erythromycin in Argentina, France, Lebanon and Spain potentially selected for GPSC10 while serotype replacement in low antibiotic consumption settings was mainly observed to be mediated by susceptible lineages.^12, 59^ Among countries with serotype 24F increase, Japan has the highest consumption of erythromycin and the least of penicillin, the serotype 24F driver lineage GPSC106 (CC2572) only exhibits erythromycin resistance.^16^ These observations suggested that vaccine- and antibiotic-selective pressure are shaping the post-vaccine population structure, and gradually the reduction in antibiotic resistance achieved by PCVs may diminish.^53^ However, resistance alone was not necessary for clonal success, as GPSC10-24F was detected in all six countries but was only expanded in high antibiotic use settings. This finding echoed the observation in other bacterial species such as *Escherichia coli*.^60^ As acquired antibiotic-resistant genes are part of the accessory genome (genes not present in all isolates of a species), their frequencies are under negative-frequency dependent selection,^61^ which may explain the coexistence of susceptible and resistant strains in the pneumococcal population.

The strength of this study lies in a large global collection of pneumococcal genomes, along with a comprehensive epidemiological metadata. Although the sampling strategy and collection time frame was not consistent between countries, the GPS project has been formulated to create pre- and post-PCV dataset in each participating country to evaluate the impact of PCV on pneumococcal population. This overarching study is complemented with the knowledge we gained from a number of country-specific analyses to provide an international perspective of GPSC10 and to highlight this lineage as a future risk in pneumococcal disease prevention. This study also underlines the need for policymakers to evaluate the overall impact of PCVs including changes in IPD incidence and detection of emerging non-vaccine serotypes. Focusing only on vaccine serotypes in estimating impact could be misleading as it would identify a rise in serotype 19A, observed in some settings with PCV10 use, but would not detect the increase in serotype 24F, associated with PCV13 use, as noted in the current study. Knowledge gained through implementation of an effective and sustainable surveillance system for *S. pneumoniae* could guide timely policy making including choice of PCV to stabilise the incidence of IPD cases at a low level or even further reduce incidence.

Our work further shows the usefulness of bacterial genome sequencing to better understand the pneumococcal lineages behind the serotype changes and reveals that GPSC10 alone is a challenge for a serotype-based vaccine strategy. More systematic investigation to identify lineages like GPSC10 will better inform and improve next-generation preventive strategies against pneumococcal disease.

## Data Availability

All data produced in the present study are available upon reasonable request to the authors

## Acknowledgement

The study was co-funded by the Bill and Melinda Gates Foundation (grant code OPP1034556) and the Wellcome Sanger Institute (core Wellcome grants 098051 and 206194). Particular thanks go to all members of the Global Pneumococcal Sequencing (GPS) Consortium for their contributions of sample collection, processing and collaborative spirit. We are grateful for feedback from Dr Adam Cohen and Dr Xin Liu from the Centers for Disease Control and Prevention and Dr Chrispin Chaguza from Yale University. We acknowledge Dr Corinne Lévy, Dr Naïm Ouldali, and Stéphane Béchet (ACTIV) who manage children data collection and have help in establishing the study sample for France. We are also grateful to Assiya El Mniai and Cécile Culeux (French NRL for pneumococci) for their technical assistance in preparing DNA for whole genome sequencing. We acknowledge the support from the sequencing facility and the Pathogen Informatics team at the Wellcome Sanger Institute The findings and conclusions detailed in this manuscript are those of the authors and do not necessarily represent the official position of the Centers for Disease Control and Prevention. For the purpose of open access, the author has applied a CC BY public copyright license to any Author Accepted Manuscript version arising from this submission.

## The Global Pneumococcal Sequencing Project Consortium

Abdullah W Brooks, Alejandra Corso, Alexander Davydov, Alison Maguire, Anmol Kiran, Benild Moiane, Bernard Beall, Chunjiang Zhao, David Aanensen, Dean B Everett, Diego Faccone, Ebenezer Foster-Nyarko, Ebrima Bojang, Ekaterina Egorova, Elena Voropaeva, Eric Sampane-Donkor, Ewa Sadowy, Geetha Nagaraj, Helio Mucavele, Houria Belabbès, Idrissa Diawara, Jennifer Verani, Jeremy Keenan, John A Lees, Jyothish N Nair Thulasee Bhai, Kedibone Ndlangisa, Khalid Zerouali, Leon Bentley, Leonid Titov, Linda De Gouveia, Maaike Alaerts, Margaret Ip, Maria Cristina de Cunto Brandileone, Md Hasanuzzaman, Metka Paragi, Michele Nurse-Lucas, Mignon du Plessis, Mushal Ali, Nicholas Croucher, Nicole Wolter, Noga Givon-Lavi, Nurit Porat, Özgen Köseoglu Eser, Pak Leung Ho, Patrick Eberechi Akpaka, Paula Gagetti, Peggy-Estelle Tientcheu, Pierra Law, Rachel Benisty, Rafal Mostowy, Roly Malaker, Samanta Cristine Grassi Almeida, Sanjay Doiphode, Shabir A. Madhi, Shamala Devi Sekaran, Stuart C Clarke, Somporn Srifuengfung, Susan A Nzenze, Tamara Kastrin, Theresa J. Ochoa, Waleria Hryniewicz, Yulia Urban

## Research in context

### Evidence before this study

We searched PubMed using the terms “streptococcus pneumoniae” AND “24F” OR “CC230” OR “GPSC10” for papers published in English between Jan 1 2000 and Feb 19 2021. We searched for population-based studies which reported changes in serotype 24F before and after the introduction of pneumococcal conjugate vaccine in the country or region. After reviewing 59 articles, 28 met the inclusion criteria. The effects of 7-valent PCV were measured in 6 studies and 13-valent PCV (PCV13) in 23 studies. The majority of studies utilised samples from children and/or adults with IPD (n=20), four studies included isolates from both carriage and IPD cases and four studies analysed isolates from carriage alone.

Studies were conducted at the national or regional level, and typed isolates using Quellung and/or latex agglutination and/or PCR based methods. Amongst IPD cases in children, 24F was identified as the predominant serotype post PCV13 in eight studies representing cases from France, Denmark, Spain, Italy and Japan. For the three studies with serotype stratified by child age, the predominance of 24F was only observed in children up to 5 years of age. Serotype 24F was the second most common serotype, or jointly the most common with 12F, in four further studies of IPD cases post PCV13 in Germany, France and the UK. An increase in 24F post PCV7 was reported amongst IPD cases from Spain, Italy and France, carriage and IPD cases in Norway and carriage in Portugal.

### Added value of this study

We have an enhanced understanding of the multidrug resistant lineage of *S. pneumoniae* which has driven the increase in serotype 24F in France post PCV13. Additionally, we utilised a global collection of *S. pneumoniae* isolates from 56 countries to contextualise the isolates from France, identifying the predominant lineage (GPSC10) associated with the increase of multidrug resistance in serotype 24F globally. This study is complemented with the knowledge we gained from a number of country-specific analyses to demonstrate GPSC10 may pose a global threat after PCV13 due to a high risk of vaccine evasion.

### Implications of all the available evidence

The increase in serotype 24F post PCV13 in France was attributed to the multidrug resistant lineage GPSC10. Concerningly, GPSC10 has a relatively high disease potential and propensity to cause meningitis independent of serotype. GPSC10 appears to be highly capable of acquiring DNA that may result in antimicrobial resistance and serotype switches. Analyses of GPSC10 isolates from a global dataset of *S. pneumoniae* genomes have identified expression of an additional16 serotypes of which only six are included in PCV13. Antimicrobial use may have contributed to selection of GPSC10 in France and Spain, decreasing the benefit of PCV for reduction of AMR. GPSC10 has transmitted amongst European countries, with long-range transmissions to other continents. The evidence suggests that the expansion of GPSC10 may be a challenging problem to address using a serotype-based vaccine strategy.

---

**Figure S1.**
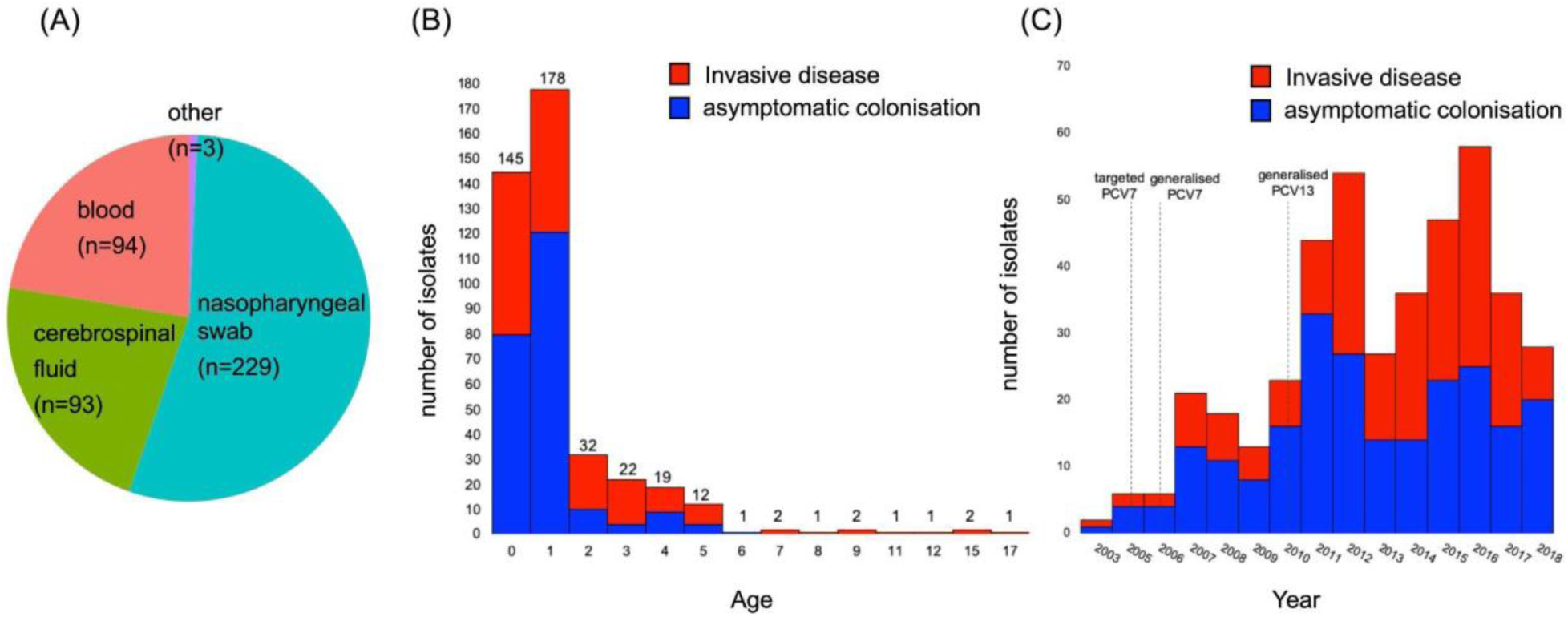
A collection of serotype 24F *Streptococcus pneumoniae* from France 2003-2018 by (A) clinical sample source, (B) age and (C) year of collection. The collection indicates an almost 1:1 ratio of samples from invasive disease (cerebrospinal fluid, blood, and others) and asymptomatic colonisation (nasopharyngeal swab) overall and over the year. Majority of samples are from children aged 2 and under.

**Figure S2.**
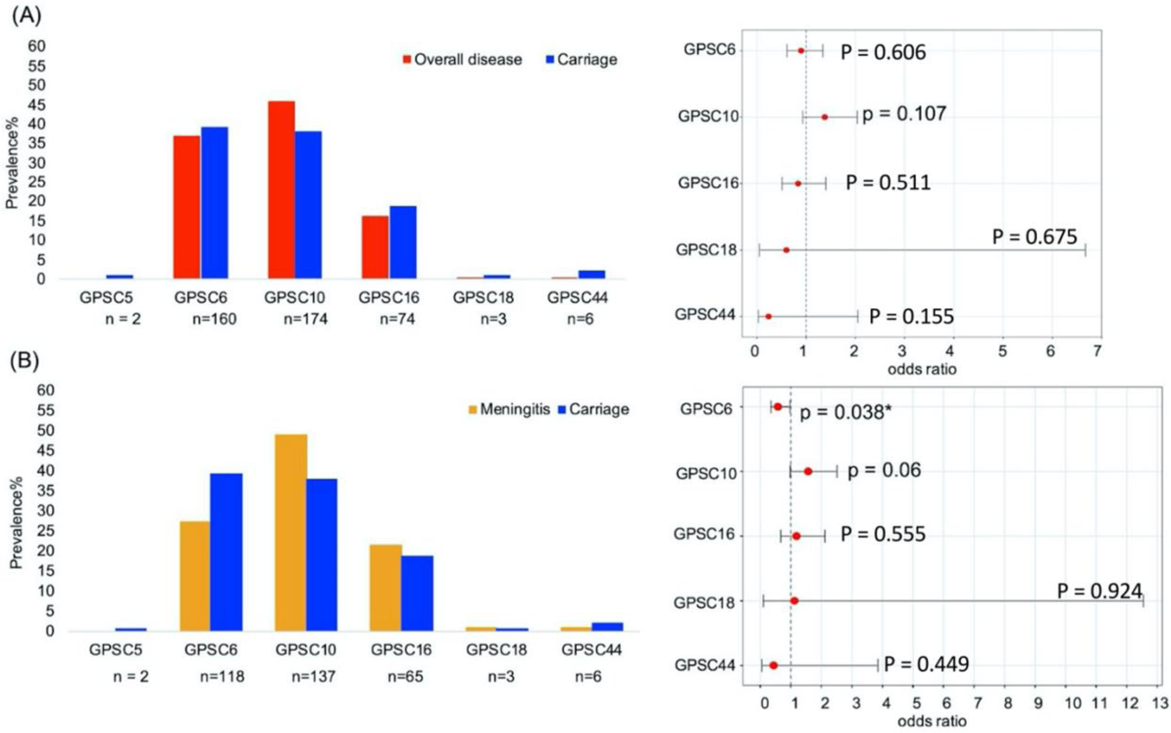
Prevalence of pneumococcal lineages by clinical manifestations and odds ratio for causing (A) overall invasive diseases and (B) meningitis by reference to carriage. The odds ratio and 95% confidence interval were calculated using Fisher’s Exact test.

**Figure S3.**
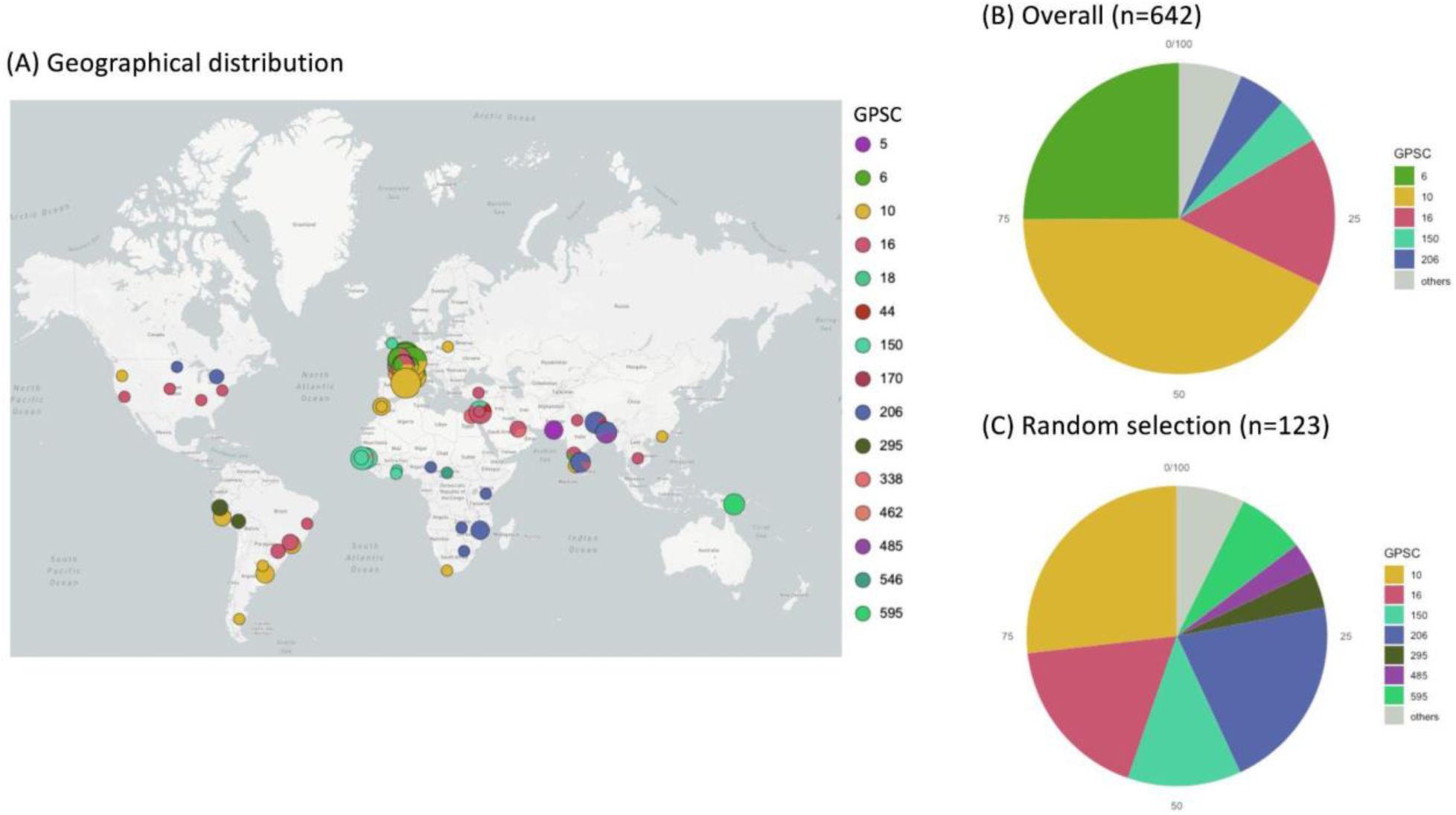
(A)The geographical distribution of serotype 24F *Streptococcus pneumoniae* (n=642) in the Global Pneumococcal Sequencing (GPS) project database, including 419 isolates from France. (B) Proportion of pneumococcal lineages or Global Pneumococcal Sequence Clusters (GPSCs) in an overall collection of serotype 24F pneumococci (n=642) and a sub-collection (n=123) includes isolates randomly selected from disease surveillance systems and carriage surveys. Pneumococcal lineages less than 3% in prevalence are grouped as others in the pie charts. The geographical distribution can be interactively visualised at https://microreact.org/project/global_24F/7d36573f.

**Figure S4.**
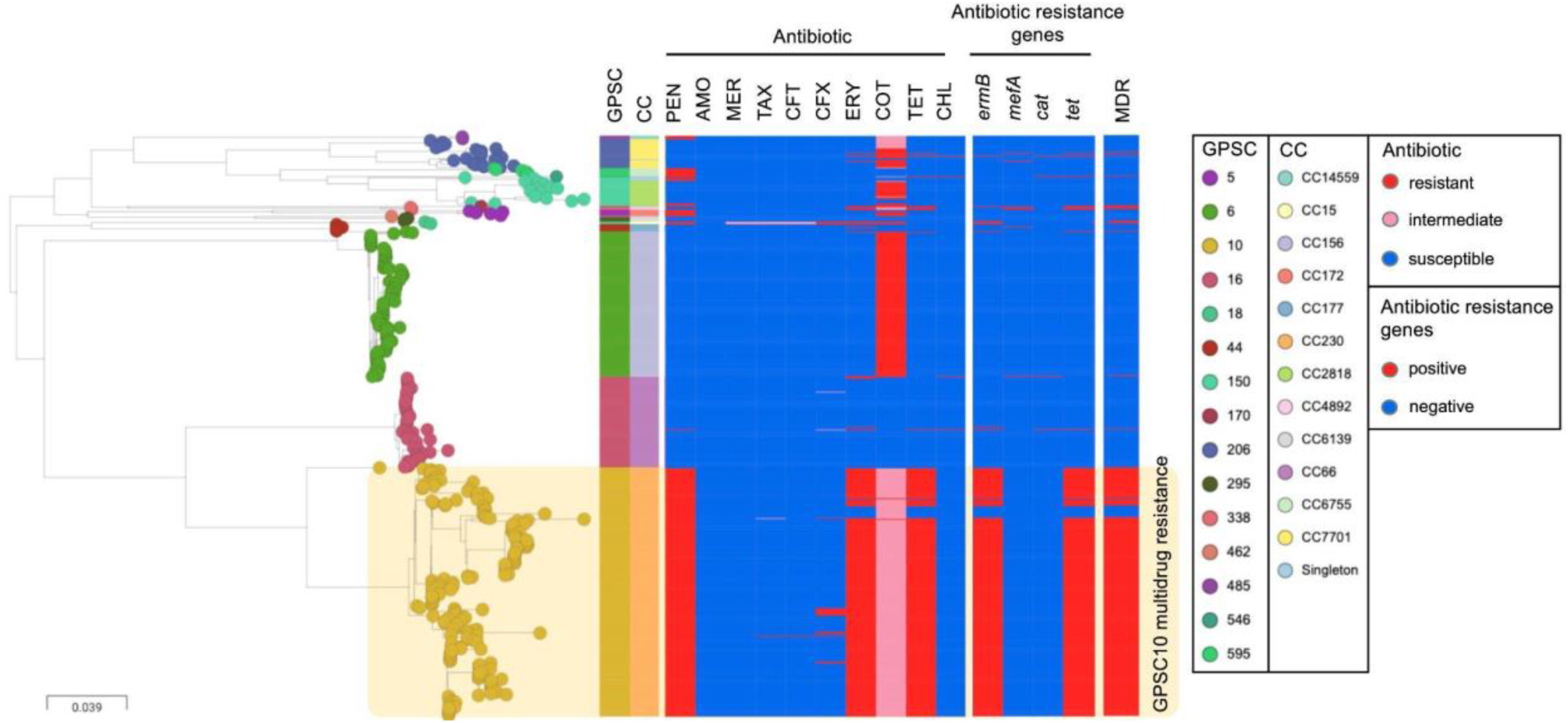
Phylogeny of 642 serotype 24F *Streptococcus pneumoniae* from 29 countries across six continents overlaid with antibiotic resistance profiles. GPSC, global pneumococcal sequence cluster; CC, clonal complex; PEN, penicillin; AMO, amoxicillin; MER, meropenem; TAX, cefotaxime; CFT, ceftriaxone; CFX, cefuroxime; ERY, erythromycin; COT, cotrimoxazole; TET, tetracycline; CHL, chloramphenicol; MDR, multidrug resistance. This figure can be interactively visualised at https://microreact.org/project/global_24F/e1acf229.

**Figure S5.**
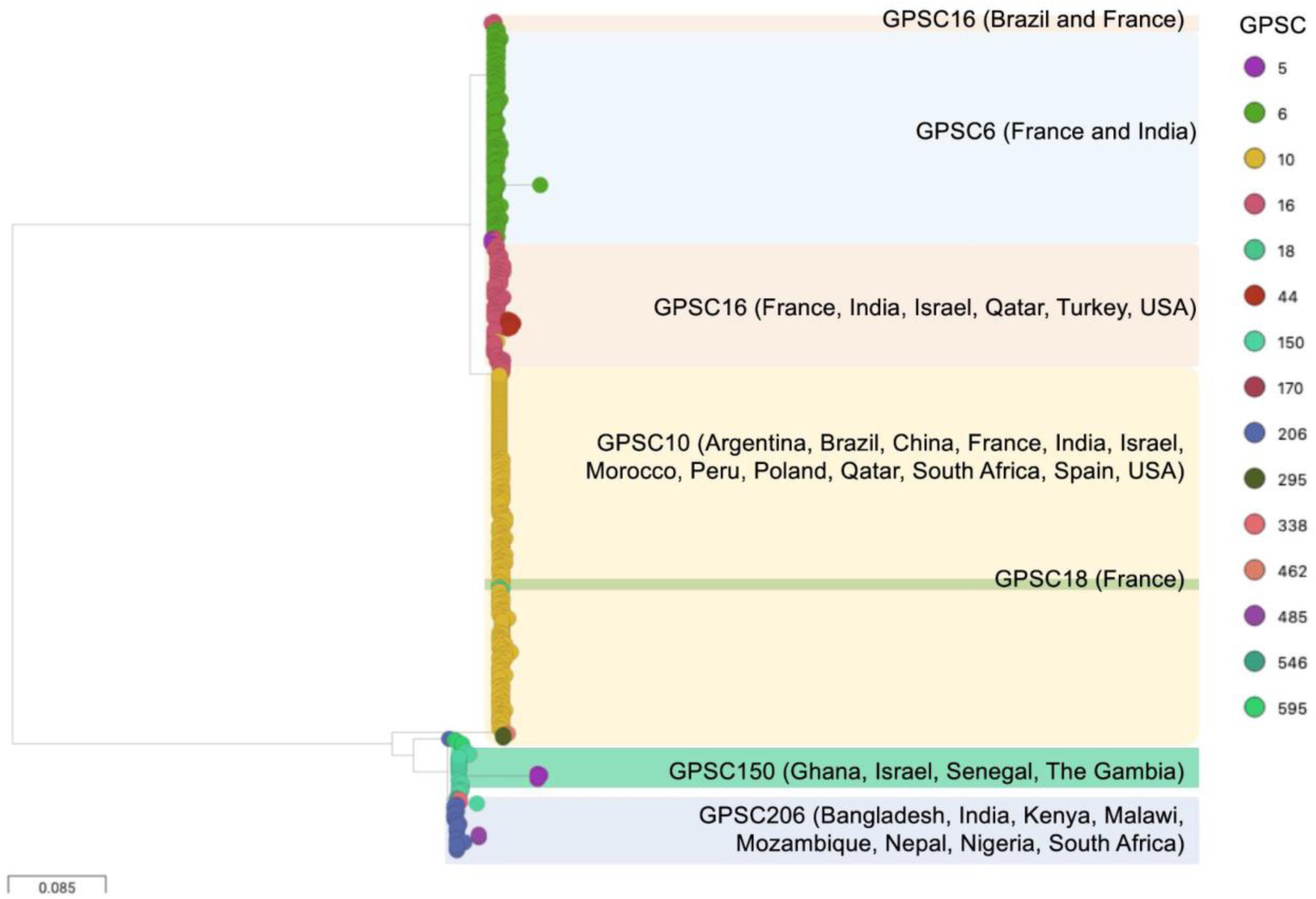
A phylogeny built upon the genetic variants identified from the capsular encoding region (*cps*) in a collection of 642 serotype 24F *Streptococcus pneumoniae* and overlaid with Global Pneumococcal Sequence Clusters (GPSCs)

**Figure S6.**
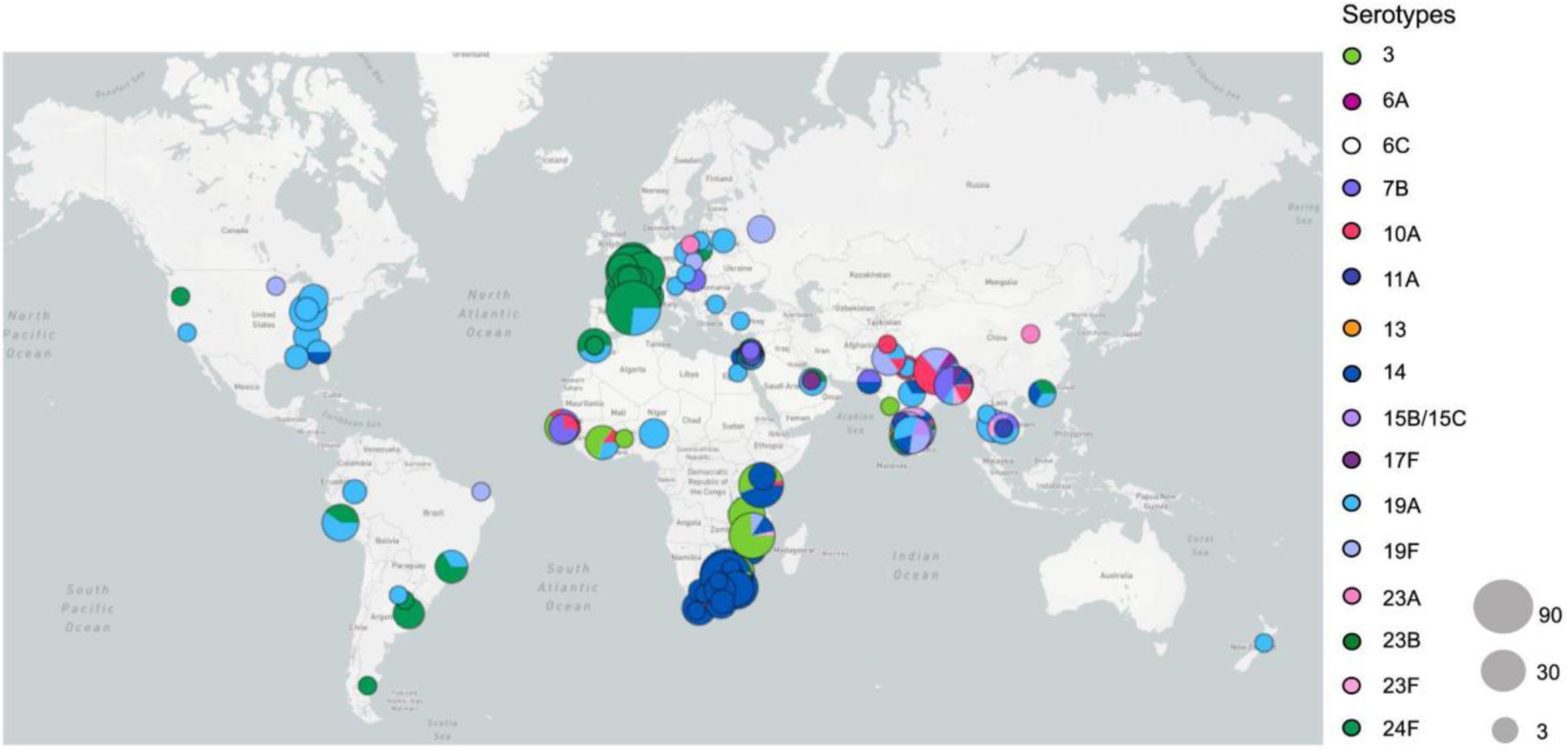
Geographical distribution of Global Pneumococcal Sequence Cluster (GPSC)10 (n=888) from 33 countries. This figure can be interactively viewed at https://microreact.org/project/global_GPSC10/21948517

**Figure S7.**
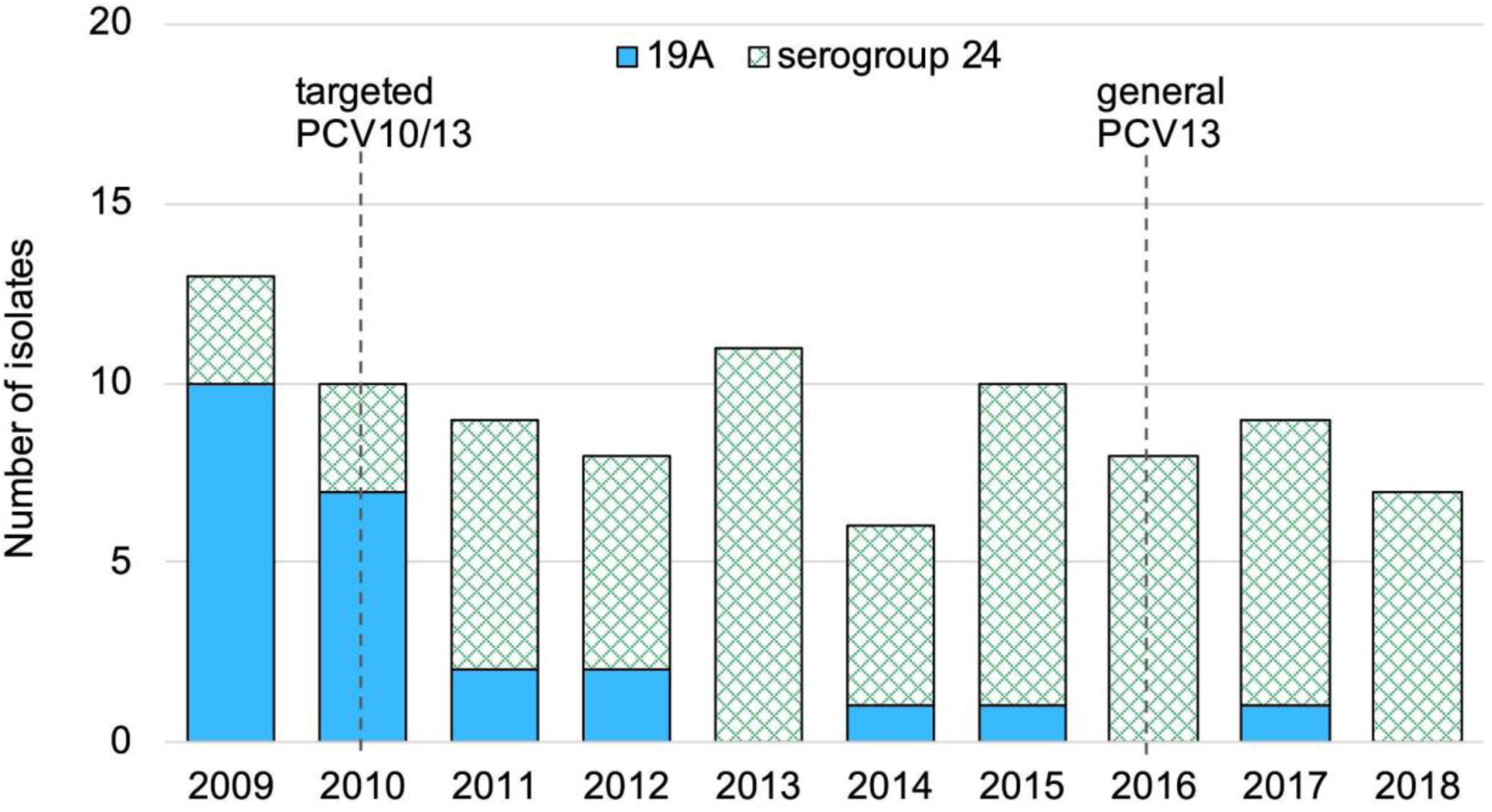
Rapid changes in serotype composition within Global Pneumococcal Sequence Cluster (GPSC)10 during PCV introductions among 91 isolates from Spain. Serotype 19A is targeted by PCV13 but not PCV10. Serotype 24F is not targeted by either.

**Figure S8.**
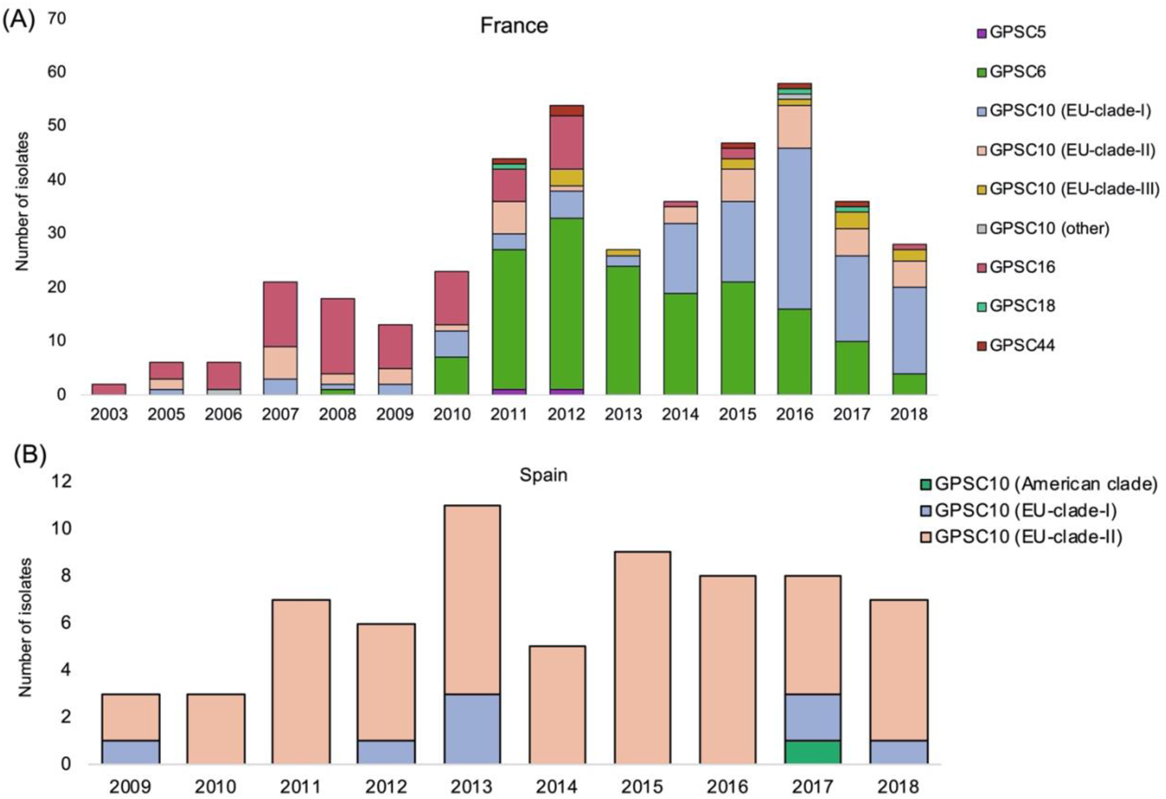
The proportion of Global Pneumococcal Sequencing Cluster (GPSC)10 clades and other GPSCs in serotype 24F *Streptococcus pneumoniae* isolates causing invasive pneumococcal disease from France (A) and Spain (B).

**Table S1.** Prevalence of GPSCs in serotype 24F *Streptococcus pneumoniae* causing invasive disease (n=190) and asymptomatic colonisation (n=229) in France over vaccine periods.

|  | n(%) |  |  |  | P value |  |  |
| --- | --- | --- | --- | --- | --- | --- | --- |
| Lineages | Targeted PCV7 (2003-2005) | Generalised PCV7 (2006-2010) | Early PCV13 (2011-2014) | Late PCV13 (2015-2018) | Targeted PCV7 vs generalised | Generalised PCV7 vs Early PCV13 | Early PCV13 vs Late PCV13 |
| GPSC5 | - | - | 2 (1.2) | - | - | - | - |
| GPSC6 | - | 8 (9.9) | 101 (62.7) | 51 (30.2) | - | <0.0001* | <0.0001* |
| GPSC10 | 3 (37.5) | 24 (29.6) | 37 (23.0) | 110 (65.1) | 1.0000 | 0.6289 | <0.0001* |
| GPSC16 | 5 (62.5) | 49 (60.5) | 17 (10.6) | 3 (1.8) | 1.0000 | <0.0001* | 0.0023* |
| GPSC18 | - | - | 1 (0.6) | 2 (1.2) | - | - | 1.0000 |
| GPSC44 | - | - | 3 (1.9) | 3 (1.8) | - | - | 1.0000 |

**Table S2.** Odds ratio for invasiveness and propensity to cause meningitis of six pneumococcal lineages expressing serotype 24F from France.

|  | % (n) |  |  | Overall disease vs carriage |  | Meningitis vs Carriage |  |
| --- | --- | --- | --- | --- | --- | --- | --- |
| GPSC | Overall disease (n=190) | Meningitis (n=102) | Carriage (n=229) | Odds ratio | P value | Odds ratio | P value |
| GPSC5 | 0 | 0 | 2 | 0 (0-Inf) | 0.197 | 0 (0-Inf) | 0.344 |
| GPSC6 | 37% (70) | 27% (28) | 39% (90) | 0.90 (0.61-1.34) | 0.606 | 0.58 (0.35-0.97) | 0.038 |
| GPSC10 | 46% (87) | 49% (50) | 38% (87) | 1.38 (0.93-2.04) | 0.107 | 1.57 (0.98-2.51) | 0.06 |
| GPSC16 | 16% (31) | 22% (22) | 19% (43) | 0.84 (0.51-1.40) | 0.511 | 1.19 (0.67-2.12) | 0.555 |
| GPSC18 | 0.5% (1) | 1% (1) | 0.9% (2) | 0.60 (0.05, 6.67) | 0.675 | 1.12 (0.10-12.54) | 0.924 |
| GPSC44 | 0.5% (1) | 1% (1) | 2% (5) | 0.24 (0.03, 2.05) | 0.155 | 0.44 (0.05-3.85) | 0.449 |

**Table S3.** A pairwise odds ratio for two samples being diverged from time-to-most-recent- common-ancestor (tMRCA) and recovered from the same French province.

| Divergence time window (tMRCA in years)<sup>a</sup> | Median divergence time window (tMRCA in years) | Odds ratio | lower 95% confidence interval | upper 95% confidence interval |
| --- | --- | --- | --- | --- |
| 0 - 2 | 1 | 5.1059 | 2.5373 | 9.5085 |
| 1 - 3 | 2 | 4.4514 | 2.4115 | 8.9007 |
| 2 - 4 | 3 | 2.0891 | 1.1846 | 3.4690 |
| 3 - 5 | 4 | 1.6435 | 0.9131 | 3.2532 |
| 4 - 6 | 5 | 1.0065 | 0.6424 | 1.6286 |
| 5 - 7 | 6 | 0.7315 | 0.5203 | 1.0835 |
| 6 - 8 | 7 | 0.5662 | 0.4320 | 0.7939 |
| 7 - 9 | 8 | 0.9479 | 0.6563 | 1.4437 |
| 8 - 10 | 9 | 1.6082 | 1.1058 | 2.1685 |
| 9 - 11 | 10 | 1.3436 | 0.9728 | 1.9553 |
| 10 - 12 | 11 | 1.5775 | 1.1832 | 2.1800 |
| 11 - 13 | 12 | 1.1970 | 0.9350 | 1.5283 |
| 12 - 14 | 13 | 0.8488 | 0.6187 | 1.0893 |
| 13 - 15 | 14 | 0.8482 | 0.5906 | 1.2123 |
| 14 - 16 | 15 | 0.9607 | 0.7505 | 1.3370 |
| 15 - 17 | 16 | 0.9494 | 0.5808 | 1.5549 |
| 16 - 18 | 17 | 0.4648 | 0.2962 | 0.8512 |
| 17 - 19 | 18 | 0.7504 | 0.4834 | 1.2583 |
| 18 - 20 | 19 | 1.2611 | 0.9385 | 1.8455 |

<sup>a</sup>The calculation of divergence time is based on a rolling window of two years.

**Table S4.**
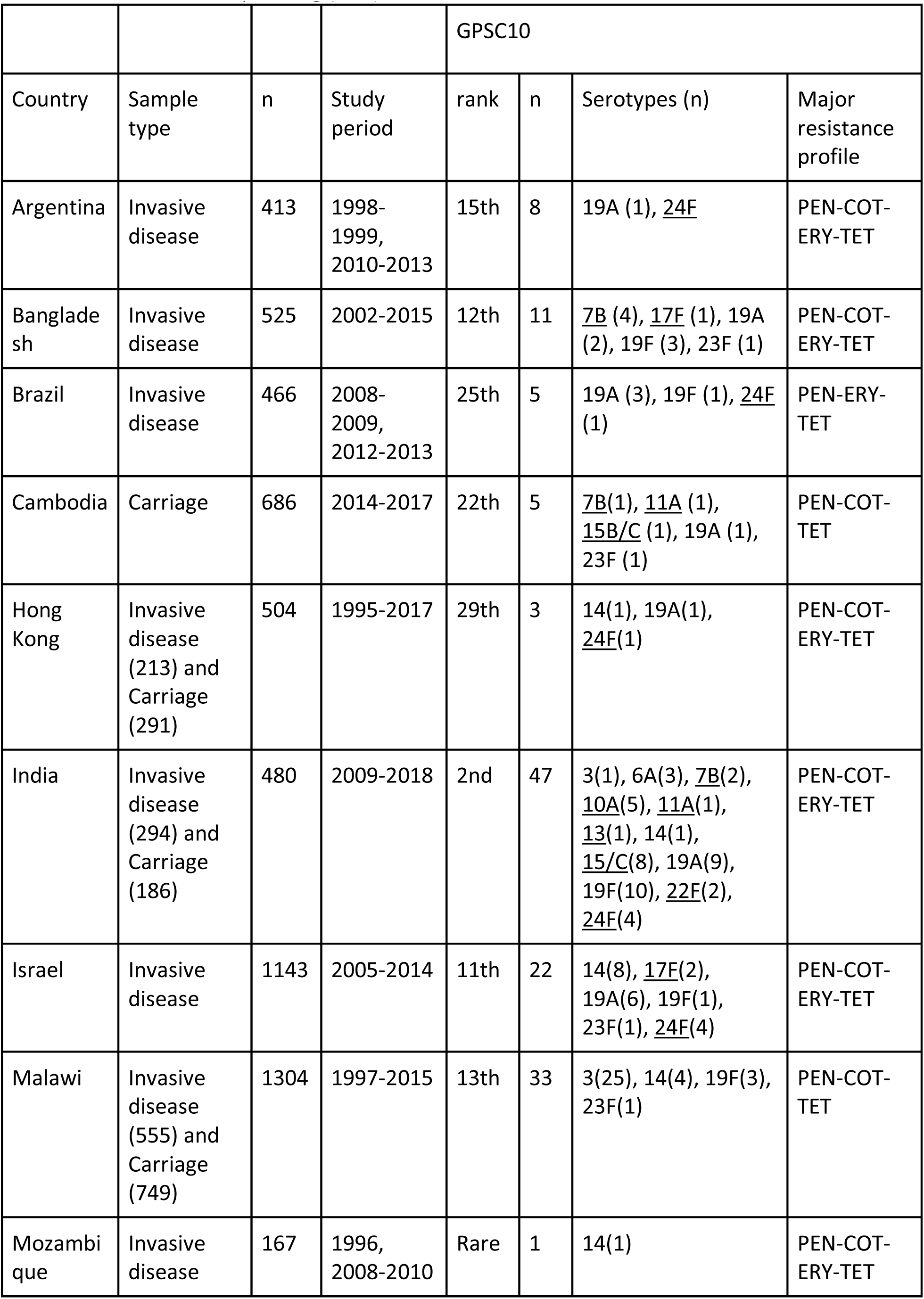
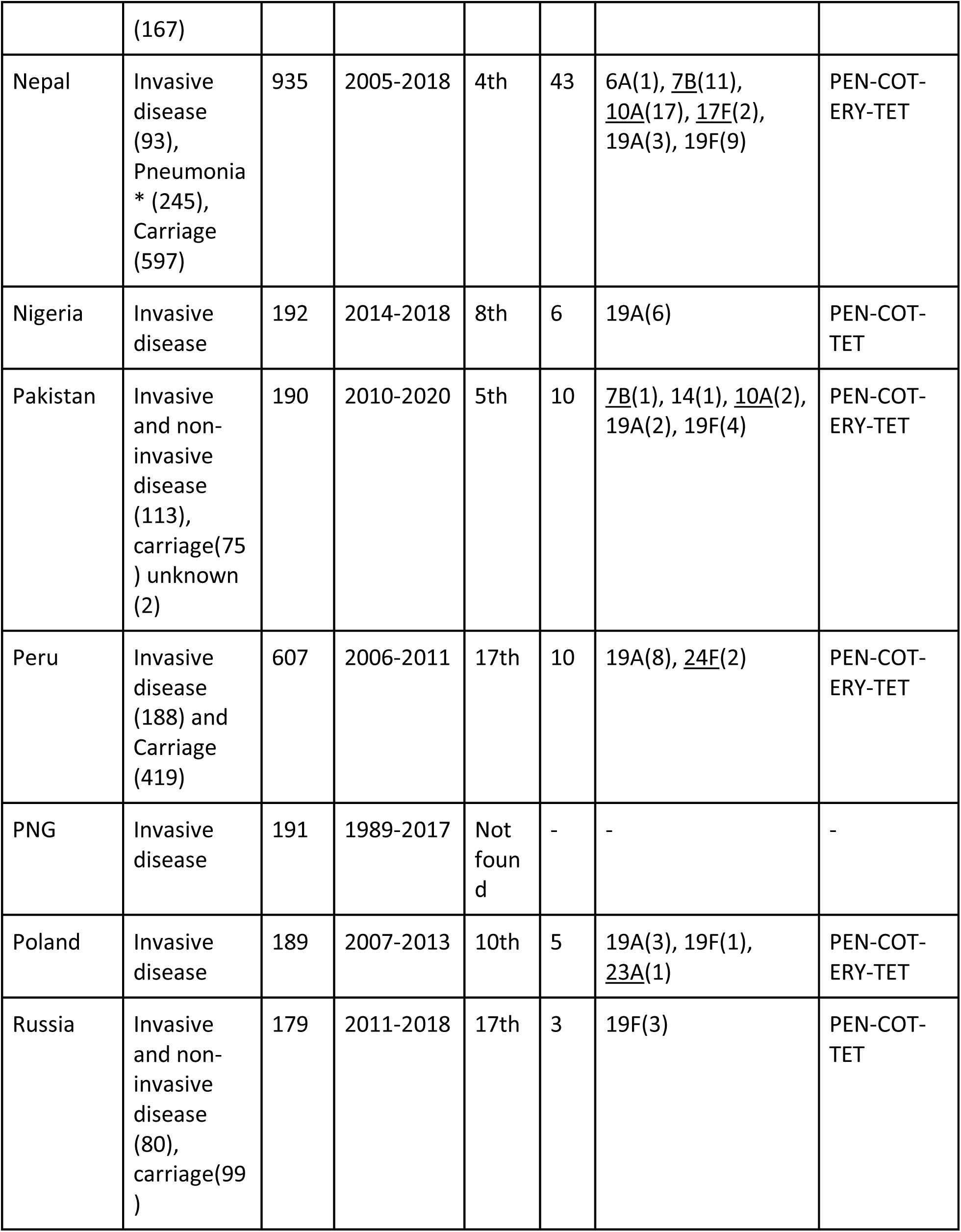
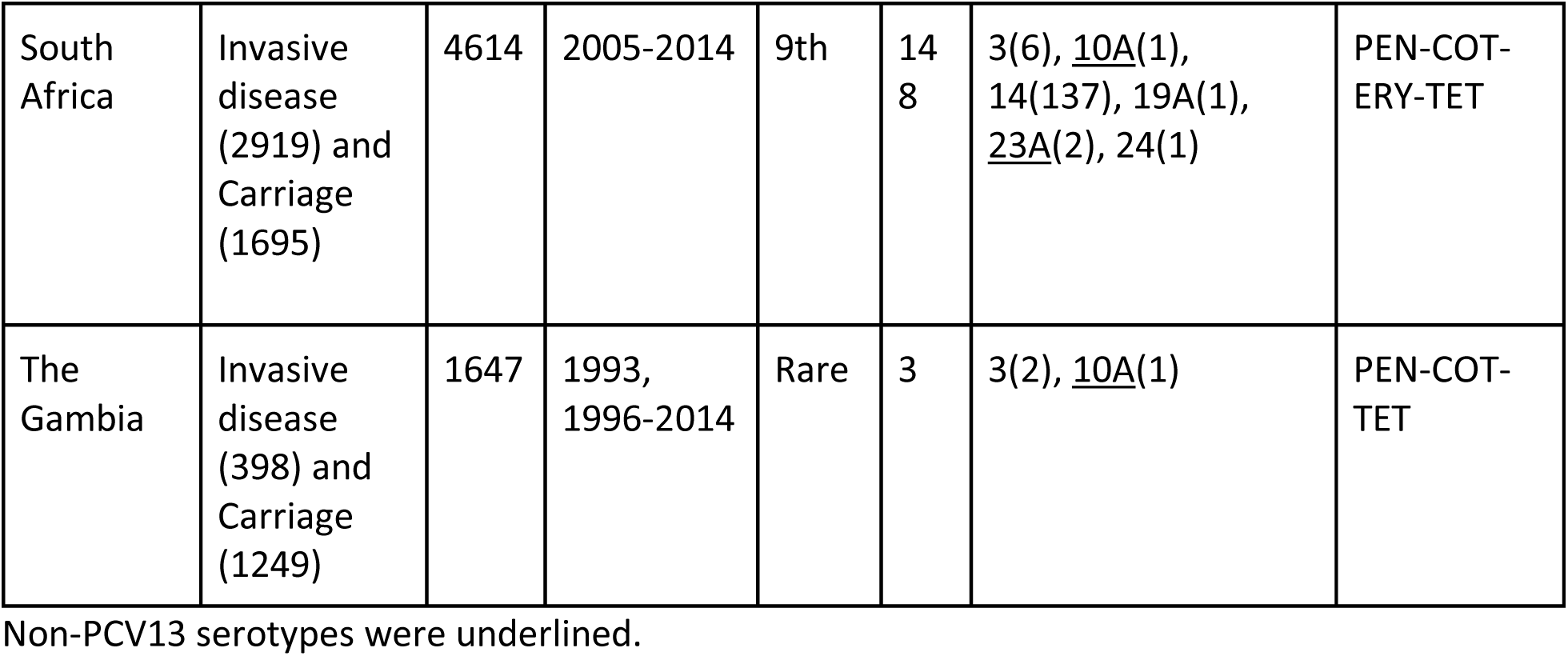
The prevalence, serotypes and resistance profile of GPSC10 by country in the Global Pneumococcal Sequencing (GPS) database

**Table S5.** The relationship between pneumococcal 24F driver lineages and antibiotic consumption.

|  | Major driver of serotype 24F |  | Narrow and broad spectrum penicillin |  | Macrolide<sup>a</sup> |  |
| --- | --- | --- | --- | --- | --- | --- |
| Countries | GPSC (CC) | Resistance profile | DDD<sup>b</sup> per 1000 | Ratio | DDD per 1000 | Ratio |
| Denmark | 6 (CC156) | COT | 4,781 | Ref | 664 | Ref |
| Argentina | 10 (CC230) | PEN-COT-ERY-TET | 4328 | 0.9 | 770 | 1.2 |
| France | 10 (CC230) | PEN-COT-ERY-TET | 8,522 | 1.8 | 1,311 | 2.0 |
| Japan | 106(CC2572) | ERY | 634 | 0.1 | 1,525 | 2.3 |
| Lebanon | 10 (CC230) | PEN-COT-ERY-TET | 6544 | 1.4 | 765 | 1.2 |
| Spain | 10 (CC230) | PEN-COT-ERY-TET | 10,389 | 2.2 | 998 | 1.5 |

Erythromycin belongs to the macrolide class of antibiotics. COT, cotrimoxazole; PEN, penicillin; ERY, erythromycin; TET, tetracycline
<sup>a</sup>Macrolide is a class of antibiotics which includes erythromycin
<sup>b</sup>DDD, defined daily dose. The DDD estimates are extracted from
https://resistancemap.cddep.org/AntibioticUse.php

## Supplementary files 2

The -80°C stock of each *S. pneumoniae* isolate was plated on an agar plate with 5% sheep blood and incubated overnight at 37°C in 5% CO2. A single colony from the overnight culture was inoculated in 5ml Todd Hewitt broth at 37°C in 5% CO2 overnight. The bacterial pellet from the overnight broth culture was then subject to DNA extraction. Pneumococcal DNA was extracted using a modified protocol of QIAamp1DNAMini Kit (QIAGEN, IncValencia, CA) protocol as previously described ^1^. The DNA quantity was evaluated by Qubit and then subject to sequencing on an Illumina HiSeq platform at Wellcome Sanger Institute, generating ≥100bp paired-end reads. The reads were assembled and annotated as previously described.^2^ Quality control of the genome sequences was as follow: 1) overall sequencing depth >20X, 2) >60% reads mapping to Streptococcus pneumoniae using Kraken, 3) >60% mapping coverage of reference genome (PMEN global clone Spain23F-1, accession number FM211187) 4) percent of heterozygous sites over total number of single nucleotide polymorphisms (SNPs) ≤ 15%, 5) total number of contigs <500 and 6) total length of the assembled genome size between 1.9- 2.3 Mb. Serotypes were predicted from the sequence reads using SeroBA.^3^ At the time of writing, SeroBA cannot differentiate serotypes within serogroup 24. Therefore, serogroup 24 isolates (n=674) in the Global Pneumococcal Sequencing (GPS) project, including those from France, identified by SeroBA were subject to phylogenetic analysis. Reads of serogroup 24 genomes were mapped to the reference sequence of 24F capsular encoding region *cps* (CR931688) using Burrows Wheeler Aligner (BWA) version 0.7.17-r1188.^4^ The alignment was then further aligned with reference sequences of 24A (CR931686) and 24B (CR931687), followed by extracting SNPs using snp-sites.^5^ A maximum likelihood tree using FastTree version 2.1.10^6^ with GTR substitution model was constructed and overlaid with phenotypic serotyping results if available. We identified 25 isolates clustered with 24A reference, 94 with serotype 24F reference, 549 in a group that 250 isolates were confirmed as 24F by the Quellung reaction in eight different laboratories, and two divergents. No isolate’s *cps* was clustered with serotype 24B reference. The 642 isolates predicted to be serotype 24F were included for further analysis.

## References

1. Wahl B, O’Brien KL, Greenbaum A, et al. Burden of Streptococcus pneumoniae and Haemophilus influenzae type b disease in children in the era of conjugate vaccines: global, regional, and national estimates for 2000–15. The Lancet Global Health 2018; 6: e744–57.

2. Mackenzie GA, Hill PC, Jeffries DJ, et al. Effect of the introduction of pneumococcal conjugate vaccination on invasive pneumococcal disease in The Gambia: a population-based surveillance study. Lancet Infect Dis 2016; 16: 703–11.

3. Savulescu C, Krizova P, Lepoutre A, et al. Effect of high-valency pneumococcal conjugate vaccines on invasive pneumococcal disease in children in SpIDnet countries: an observational multicentre study. Lancet Respir Med 2017. DOI:10.1016/S2213-2600(17)30110-8.

4. Gottberg A von, von Gottberg A, de Gouveia L, et al. Effects of Vaccination on Invasive Pneumococcal Disease in South Africa. New England Journal of Medicine. 2014; 371: 1889–99.

5. Ladhani SN, Collins S, Djennad A, et al. Rapid increase in non-vaccine serotypes causing invasive pneumococcal disease in England and Wales, 2000–17: a prospective national observational cohort study. Lancet Infect Dis 2018; 18: 441–51.

6. Ben-Shimol S, Givon-Lavi N, Grisaru-Soen G, et al. Comparative incidence dynamics and serotypes of meningitis, bacteremic pneumonia and other-IPD in young children in the PCV era: Insights from Israeli surveillance studies. Vaccine 2017; published online June 1. DOI:10.1016/j.vaccine.2017.05.059.

7. Pai R, Moore MR, Pilishvili T, et al. Postvaccine genetic structure of Streptococcus pneumoniae serotype 19A from children in the United States. J Infect Dis 2005; 192: 1988– 95.

8. Ouldali N, Levy C, Varon E, et al. Incidence of paediatric pneumococcal meningitis and emergence of new serotypes: a time-series analysis of a 16-year French national survey. Lancet Infect Dis 2018; 18: 983–91.

9. Ouldali N, Varon E, Levy C, et al. Invasive pneumococcal disease incidence in children and adults in France during the pneumococcal conjugate vaccine era: an interrupted time-series analysis of data from a 17-year national prospective surveillance study. Lancet Infect Dis 2021; 21: 137–47.

10. Paula Gagetti, Stephanie W. Lo, Paulina A. Hawkins, Rebecca A. Gladstone, Mabel Regueira, Diego Faccone, SIREVA-Argentina group, Keith P. Klugman, Robert F. Breiman, Lesley McGee, Stephen D. Bentley, Alejandra Corso. Population genetic structure, serotype distribution and antibiotic resistance of Streptococcus pneumoniae causing invasive disease in children in Argentina. Microbial Genomics (accepted).

11. Adam HJ, Golden AR, Karlowsky JA, et al. Analysis of multidrug resistance in the predominant Streptococcus pneumoniae serotypes in Canada: the SAVE study, 2011–15. J Antimicrob Chemother 2018; 73: vii12–9.

12. Kavalari ID, Fuursted K, Krogfelt KA, Slotved H-C. Molecular characterization and epidemiology of Streptococcus pneumoniae serotype 24F in Denmark. Sci Rep 2019; 9: 5481.

13. van der Linden M, Falkenhorst G, Perniciaro S, Imohl M. Effects of Infant Pneumococcal Conjugate Vaccination on Serotype Distribution in Invasive Pneumococcal Disease among Children and Adults in Germany. PLoS One 2015; 10: e0131494.

14. Phillips MT, Warren JL, Givon-Lavi N, et al. Evaluating post-vaccine expansion patterns of pneumococcal serotypes. Vaccine 2020; 38: 7756–63.

15. Camilli R, D’Ambrosio F, Del Grosso M, et al. Impact of pneumococcal conjugate vaccine (PCV7 and PCV13) on pneumococcal invasive diseases in Italian children and insight into evolution of pneumococcal population structure. Vaccine 2017; 35: 4587–93.

16. Ubukata K, Takata M, Morozumi M, et al. Effects of Pneumococcal Conjugate Vaccine on Genotypic Penicillin Resistance and Serotype Changes, Japan, 2010-2017. Emerg Infect Dis 2018; 24: 2010–20.

17. Reslan L, Finianos M, Bitar I, et al. The Emergence of Invasive Streptococcus pneumoniae Serotype 24F in Lebanon: Complete Genome Sequencing Reveals High Virulence and Antimicrobial Resistance Characteristics. Front Microbiol 2021; 12: 637813.

18. Siira L, Vestrheim DF, Winje BA, Caugant DA, Steens A. Antimicrobial susceptibility and clonality of Streptococcus pneumoniae isolates recovered from invasive disease cases during a period with changes in pneumococcal childhood vaccination, Norway, 2004–2016. Vaccine. 2020; 38: 5454–63.

19. Miguel S de, de Miguel S, Domenech M, et al. Nationwide Trends of Invasive Pneumococcal Disease in Spain From 2009 Through 2019 in Children and Adults During the Pneumococcal Conjugate Vaccine Era. Clinical Infectious Diseases. 2020. DOI:10.1093/cid/ciaa1483.

20. Latasa Zamalloa P, Sanz Moreno JC, Ordobás Gavín M, et al. Trends of invasive pneumococcal disease and its serotypes in the Autonomous Community of Madrid. Enferm Infecc Microbiol Clin 2018; 36: 612–20.

21. Kandasamy R, Voysey M, Collins S, et al. Persistent Circulation of Vaccine Serotypes and Serotype Replacement After 5 Years of Infant Immunization With 13-Valent Pneumococcal Conjugate Vaccine in the United Kingdom. J Infect Dis 2020; 221: 1361–70.

22. Silva-Costa C, Brito MJ, Aguiar SI, Lopes JP, Ramirez M, Melo-Cristino J. Dominance of vaccine serotypes in pediatric invasive pneumococcal infections in Portugal (2012–2015). Sci Rep 2019; 9: 1–9.

23. Desmet S, Wouters I, Van Heirstraeten L, et al. In-depth analysis of pneumococcal serotypes in Belgian children (2015–2018): Diversity, invasive disease potential, and antimicrobial susceptibility in carriage and disease. Vaccine. 2021; 39: 372–9.

24. Balsells E, Dagan R, Yildirim I, et al. The relative invasive disease potential of Streptococcus pneumoniae among children after PCV introduction: a systematic review and meta-analysis. J Infect 2018; published online June 29. DOI:10.1016/j.jinf.2018.06.004.

25. Gladstone RA, Lo SW, Lees JA, et al. International genomic definition of pneumococcal lineages, to contextualise disease, antibiotic resistance and vaccine impact. EBioMedicine 2019; 43: 338–46.

26. Enright MC, Spratt BG. A multilocus sequence typing scheme for Streptococcus pneumoniae: identification of clones associated with serious invasive disease. Microbiology 1998; 144: 3049–60.

27. Ludwig G, Garcia-Garcia S, Lanaspa M, et al. Serotype and clonal distribution dynamics of invasive pneumococcal strains after PCV13 introduction (2011-2016): Surveillance data from 23 sites in Catalonia, Spain. PLoS One 2020; 15: e0228612.

28. Ouldali N, Cohen R, Levy C, et al. Pneumococcal susceptibility to antibiotics in carriage: a 17 year time series analysis of the adaptive evolution of non-vaccine emerging serotypes to a new selective pressure environment. J Antimicrob Chemother 2019; 74: 3077–86.

29. Brueggemann AB, Griffiths DT, Meats E, Peto TE, Crook DW, Spratt BG. Clonal Relationships between Invasive and Carriage Streptococcus pneumoniae and Serotype- and Clone- Specific Differences in Invasive Disease Potential. J Infect Dis 2003; 187: 1424–32.

30. Lees JA, Harris SR, Tonkin-Hill G, et al. Fast and flexible bacterial genomic epidemiology with PopPUNK. Genome Res 2019; 29: 304–16.

31. J. Page A, Taylor B, A. Keane J. Multilocus sequence typing by blast from de novo assemblies against PubMLST. J Open Source Softw 2016; 1: 118.

32. Feil EJ, Li BC, Aanensen DM, Hanage WP, Spratt BG. eBURST: Inferring Patterns of Evolutionary Descent among Clusters of Related Bacterial Genotypes from Multilocus Sequence Typing Data. J Bacteriol 2004; 186: 1518–30.

33. Epping L, van Tonder AJ, Gladstone RA, et al. SeroBA: rapid high-throughput serotyping of Streptococcus pneumoniae from whole genome sequence data. Microb Genom 2018; 4. DOI:10.1099/mgen.0.000186.

34. Price MN, Dehal PS, Arkin AP. FastTree 2--approximately maximum-likelihood trees for large alignments. PLoS One 2010; 5: e9490.

35. Wellcome Trust Sanger Institute. SMALT. 2014. http://www.sanger.ac.uk/science/tools/smalt-0.

36. Li H, Durbin R. Fast and accurate short read alignment with Burrows-Wheeler transform. Bioinformatics 2009; 25: 1754–60.

37. Argimón S, Abudahab K, Goater RJE, et al. Microreact: visualizing and sharing data for genomic epidemiology and phylogeography. Microb Genom 2016; 2: e000093.

38. Croucher NJ, Page AJ, Connor TR, et al. Rapid phylogenetic analysis of large samples of recombinant bacterial whole genome sequences using Gubbins. Nucleic Acids Res 2015; 43: e15.

39. Stamatakis A. RAxML-VI-HPC: maximum likelihood-based phylogenetic analyses with thousands of taxa and mixed models. Bioinformatics 2006; 22: 2688–90.

40. Bouckaert R, Heled J, Kühnert D, et al. BEAST 2: a software platform for Bayesian evolutionary analysis. PLoS Comput Biol 2014; 10: e1003537.

41. Salje H, Lessler J, Maljkovic Berry I, et al. Dengue diversity across spatial and temporal scales: Local structure and the effect of host population size. Science 2017; 355: 1302–6.

42. Benjamini Y, Hochberg Y. Controlling the false discovery rate: A practical and powerful approach to multiple testing. J R Stat Soc 1995; 57: 289–300.

43. Nagaraj G, Govindan V, Ganaie F, et al. Streptococcus pneumoniae genomic datasets from an Indian population describing pre-vaccine evolutionary epidemiology using a whole genome sequencing approach. Microb Genom 2021; 7. DOI:10.1099/mgen.0.000645.

44. Lo SW, Gladstone RA, van Tonder AJ, et al. A mosaic tetracycline resistance gene tet(S/M) detected in an MDR pneumococcal CC230 lineage that underwent capsular switching in South Africa. J Antimicrob Chemother 2020; 75: 512–20.

45. Simoes AS, Pereira L, Nunes S, Brito-Avô A, de Lencastre H, Sa-Leao R. Clonal Evolution Leading to Maintenance of Antibiotic Resistance Rates among Colonizing Pneumococci in the PCV7 Era in Portugal. J OURNAL OF C LINICAL M ICROBIOLOGY 2011; 49: 2810–7.

46. Gherardi G, D’Ambrosio F, Visaggio D, Dicuonzo G, Del Grosso M, Pantosti A. Serotype and clonal evolution of penicillin-nonsusceptible invasive Streptococcus pneumoniae in the 7-valent pneumococcal conjugate vaccine era in Italy. Antimicrob Agents Chemother 2012; 56: 4965–8.

47. Setchanova LP, Alexandrova A, Dacheva D, Mitov I, Kaneva R, Mitev V. Dominance of multidrug-resistant Denmark(14)-32 (ST230) clone among Streptococcus pneumoniae serotype 19A isolates causing pneumococcal disease in Bulgaria from 1992 to 2013. Microb Drug Resist 2015; 21: 35–42.

48. Ardanuy C, Marimón JM, Calatayud L, et al. Epidemiology of invasive pneumococcal disease in older people in Spain (2007-2009): implications for future vaccination strategies. PLoS One 2012; 7: e43619.

49. Mayanskiy N, Savinova T, Alyabieva N, et al. Antimicrobial resistance, penicillin-binding protein sequences, and pilus islet carriage in relation to clonal evolution of Streptococcus pneumoniae serotype 19A in Russia, 2002-2013. Epidemiol Infect 2017; 145: 1708–19.

50. Hurmic O, Grall N, Al Nakib M, et al. Evidence of a clonal expansion of Streptococcus pneumoniae serotype 19A in adults as in children assessed by the DiversiLab® system. Eur J Clin Microbiol Infect Dis 2014; 33: 2067–73.

51. Muñoz-Almagro C, Esteva C, de Sevilla MF, Selva L, Gene A, Pallares R. Emergence of invasive pneumococcal disease caused by multidrug-resistant serotype 19A among children in Barcelona. J Infect 2009; 59: 75–82.

52. Balsells E, Guillot L, Nair H, Kyaw MH. Serotype distribution of Streptococcus pneumoniae causing invasive disease in children in the post-PCV era: A systematic review and meta-analysis. PLoS One 2017; 12: e0177113.

53. Lo SW, Gladstone RA, van Tonder AJ, et al. Pneumococcal lineages associated with serotype replacement and antibiotic resistance in childhood invasive pneumococcal disease in the post-PCV13 era: an international whole-genome sequencing study. Lancet Infect Dis 2019; 19: 759–69.

54. Chi-Jen L. Bacterial capsular polysaccharides—biochemistry, immunity and vaccine. Mol Immunol 1987; 24: 1005–19.

55. Isolate information: id-153 (M132-14) - Streptococcus pneumoniae isolates. https://pubmlst.org/bigsdb?page=info&db=pubmlst_spneumoniae_isolates&id=153 (accessed Sept 28, 2021).

56. Pantosti A, Gherardi G, Conte M, Faella F, Dicuonzo G, Beall B. A Novel, Multiple Drug– Resistant, Serotype 24F Strain of Streptococcus pneumoniae That Caused Meningitis in Patients in Naples, Italy. Clin Infect Dis 2002; 35: 205–8.

57. Horácio AN, Silva-Costa C, Diamantino-Miranda J, et al. Population Structure of Streptococcus pneumoniae Causing Invasive Disease in Adults in Portugal before PCV13 Availability for Adults: 2008-2011. PLoS One 2016; 11: e0153602.

58. González-Díaz A, Càmara J, Ercibengoa M, et al. Emerging non-13-valent pneumococcal conjugate vaccine (PCV13) serotypes causing adult invasive pneumococcal disease in the late-PCV13 period in Spain. Clin Microbiol Infect 2020; 26: 753–9.

59. Vestrheim DF, Steinbakk M, Aaberge IS, Caugant DA. Postvaccination increase in serotype 19A pneumococcal disease in Norway is driven by expansion of penicillin-susceptible strains of the ST199 complex. Clin Vaccine Immunol 2012; 19: 443–5.

60. Gladstone RA, McNally A, Pöntinen AK, et al. Emergence and dissemination of antimicrobial resistance in Escherichia coli causing bloodstream infections in Norway in 2002–17: a nationwide, longitudinal, microbial population genomic study. The Lancet Microbe. 2021; 2: e331–41.

61. Corander J, Fraser C, Gutmann MU, et al. Frequency-dependent selection in vaccine-associated pneumococcal population dynamics. Nat Ecol Evol 2017; 1: 1950–60.

## References

1. Hawkins PA, Akpaka PE, Nurse-Lucas M, et al. Antimicrobial resistance determinants and susceptibility profiles of pneumococcal isolates recovered in Trinidad and Tobago. Journal of Global Antimicrobial Resistance 2017; published online Aug 14. DOI:10.1016/j.jgar.2017.08.004.

2. Page AJ, De Silva N, Hunt M, et al. Robust high-throughput prokaryote de novo assembly and improvement pipeline for Illumina data. Microb Genom 2016; 2: e000083.

3. Epping L, van Tonder AJ, Gladstone RA, et al. SeroBA: rapid high-throughput serotyping of Streptococcus pneumoniae from whole genome sequence data. Microb Genom 2018; 4. DOI:10.1099/mgen.0.000186.

4. Li H, Durbin R. Fast and accurate short read alignment with Burrows-Wheeler transform. Bioinformatics 2009; 25: 1754–60.

5. Page AJ, Taylor B, Delaney AJ, et al.SNP-sites: rapid efficient extraction of SNPs from multi-FASTA alignments. Microb Genom 2016; 2: e000056.

6. Price MN, Dehal PS, Arkin AP. FastTree 2--approximately maximum-likelihood trees for large alignments. PLoS One 2010; 5: e9490.

